# Clinical evaluation of artificial intelligence for diagnostics of antibiotic-resistant bacteria

**DOI:** 10.64898/2026.08.27.26361401

**Authors:** Magnus Hessel, Juan Salvador Inda-Díaz, Anders Sjöberg, Francisco Salvà Serra, Lisa Helldal, Mats Jirstrand, Anna Johnning, Erik Kristiansson, Susann Skovbjerg

## Abstract

Antimicrobial resistance is a public health challenge, driving the need for rapid, cost-effective diagnostic support tools. Artificial intelligence (AI) may enable prediction of susceptibility to untested antibiotics from known susceptibility results, but prospective clinical validation is required before routine use. We evaluated an AI-based decision support method, trained on invasive isolates from the European Surveillance System (TESSy), for prediction of antibiotic susceptibility in clinical *Escherichia coli* urine isolates.

The evaluation included 99 *E. coli* isolates from urine samples with diversity in age, sex, and antibiotic susceptibility. Predictions were evaluated for 14 antibiotics using patient metadata and susceptibility results for 4–8 antibiotics as input. Prediction uncertainty was handled using conformal prediction, allowing abstention when confidence was insufficient. EUCAST disk diffusion test results were used as reference and genomic sequence data was used to explore mechanisms of the AI performance.

Without conformal prediction, 84% of predictions were correct when susceptibility results of six antibiotics were used to predict susceptibility to eight additional antibiotics. Across all predictions generated using susceptibility results for six antibiotics as input, the major and very major error rates were 19% and 12%, respectively. Prediction errors varied between antibiotics and were associated with certain phenotypic and genotypic resistance patterns. Conformal prediction reduced errors but increased abstentions; at confidence levels of 90%, 95%, and 97.5%, the model abstained in 9.6%, 14%, and 22% of instances. The method showed promising performance, but its clinical use remains limited and may require diagnostic data beyond susceptibility test results and demographic variables.

**Importance:** Antibiotic susceptibility testing is essential for guiding treatment of bacterial infections, but results are often incomplete when initial treatment decisions are made. This study evaluates a novel diagnostic concept: using artificial intelligence to extend the information obtained from partial susceptibility test results, rather than replacing routine susceptibility testing. The method predicts susceptibility to untested antibiotics from patient metadata and existing phenotypic susceptibility results. In this prospective evaluation of clinical *Escherichia coli* urine isolates, we assessed both prediction performance and uncertainty control by conformal prediction, which allows the method to abstain when predictions are insufficiently reliable. By linking prediction errors to phenotypic and genotypic resistance patterns, the study identified both potential and current limitations of AI-based susceptibility prediction. These findings move AI-based antimicrobial resistance prediction from retrospective model development toward prospective clinical evaluation, a necessary step before implementation.

## Introduction

The World Health Organization (WHO) has declared antibiotic resistance one of the world’s top public health and development threats. In 2024, it was estimated that infections caused by antibiotic-resistant bacteria were associated with nearly five million deaths annually, of which more than one million were caused by bacteria resistant to available antibiotics (1).

Antibiotic resistance implies that a bacterium is unlikely to be inhibited by an antibiotic at clinically relevant concentrations. Some bacterial species exhibit intrinsic resistance to certain antibiotics, but resistance may also arise from genetic changes in individual bacterial cells. For example, bacteria can acquire genes encoding enzymes that hydrolyze beta-lactam antibiotics. Extended-spectrum beta-lactamases (ESBLs) are beta-lactamases that hydrolyze many extended-spectrum cephalosporins, particularly third-generation cephalosporins, and represent a key mechanism of clinically relevant beta-lactam resistance in *Enterobacterales*. Resistance phenotypes are often correlated across antibiotics because of shared resistance mechanisms or co-resistance resulting from the carriage of multiple resistance determinants on the same mobile genetic elements (2, 3). These correlations may be learned by artificial intelligence and used to predict susceptibility to untested antibiotics from existing susceptibility test results.

Antibiotic susceptibility testing (AST) typically assesses bacterial growth in the presence of a specific antibiotic. The resulting measurements are interpreted using standardized criteria based on clinical, pharmacological, and microbiological evidence. These criteria link test results to treatment outcomes and are used to classify isolates into susceptibility categories. Standards are maintained by organizations such as the European Committee on Antimicrobial Susceptibility Testing (EUCAST) (4) and the Clinical and Laboratory Standards Institute (CLSI) (5). Despite standardization, current diagnostic approaches have limitations in speed, workload and measurement uncertainty. Moreover, each antibiotic-isolate combination generally requires a specific measurement.

Artificial intelligence (AI) has a growing range of applications in clinical diagnostics, particularly in medical imaging (6), and is increasingly being used for antibiotic susceptibility assessment. AI-based approaches have used MALDI-TOF mass spectra, whole-genome sequencing, or metagenomic sequencing to predict antibiotic susceptibility or minimum inhibitory concentrations (MICs) for different bacterial species and antibiotics (7–13). Beyond laboratory-based approaches, AI has also been applied to clinical data, where machine learning methods support antibiotic therapy decisions and predict resistance patterns from electronic medical records (14, 15). However, the implementation of AI in routine microbiological diagnostics remains limited, and evidence for its medical benefit as decision support is still scarce. A necessary step toward such evidence is prospective evaluation of AI methods (16).

In this prospective study, we evaluated an AI method for predicting whether a bacterial isolate is susceptible or resistant to an antibiotic based on partial diagnostic data (17). The method is based on neural networks using transformer architecture, and uses partial diagnostic data as input, including age, sex, country, sampling date, and available phenotypic susceptibility test results for at least four antibiotics (17). Although the AI method performs well on reference data (17), clinical validation is critical before it can be deployed in routine diagnostics. This study aimed to prospectively evaluate the performance of the AI-based decision support method for predicting antibiotic susceptibility in clinical *E. coli* isolates from patient urine samples and to investigate patterns of incorrect predictions. Overall, the method showed promising predictive performance, although error rates varied between antibiotics, were higher for susceptible isolates, and were associated with specific phenotypic and genotypic resistance patterns.

## Materials and methods

### Clinical evaluation setup

The evaluation was based on *E. coli* isolates selected from routine urine cultures that underwent antibiotic susceptibility testing at the Department of Clinical Microbiology of Sahlgrenska University Hospital, Gothenburg, Sweden (Fig. 1). Input diagnostic data to the AI method consisted of age, sex, country, sampling date and dichotomized susceptibility test results for 4–8 antibiotics, and the output was predicted susceptibility to the remaining antibiotics. Predicted susceptibility categories were compared with the measured AST results, and the performance of the AI method was calculated.

**Figure 1.**
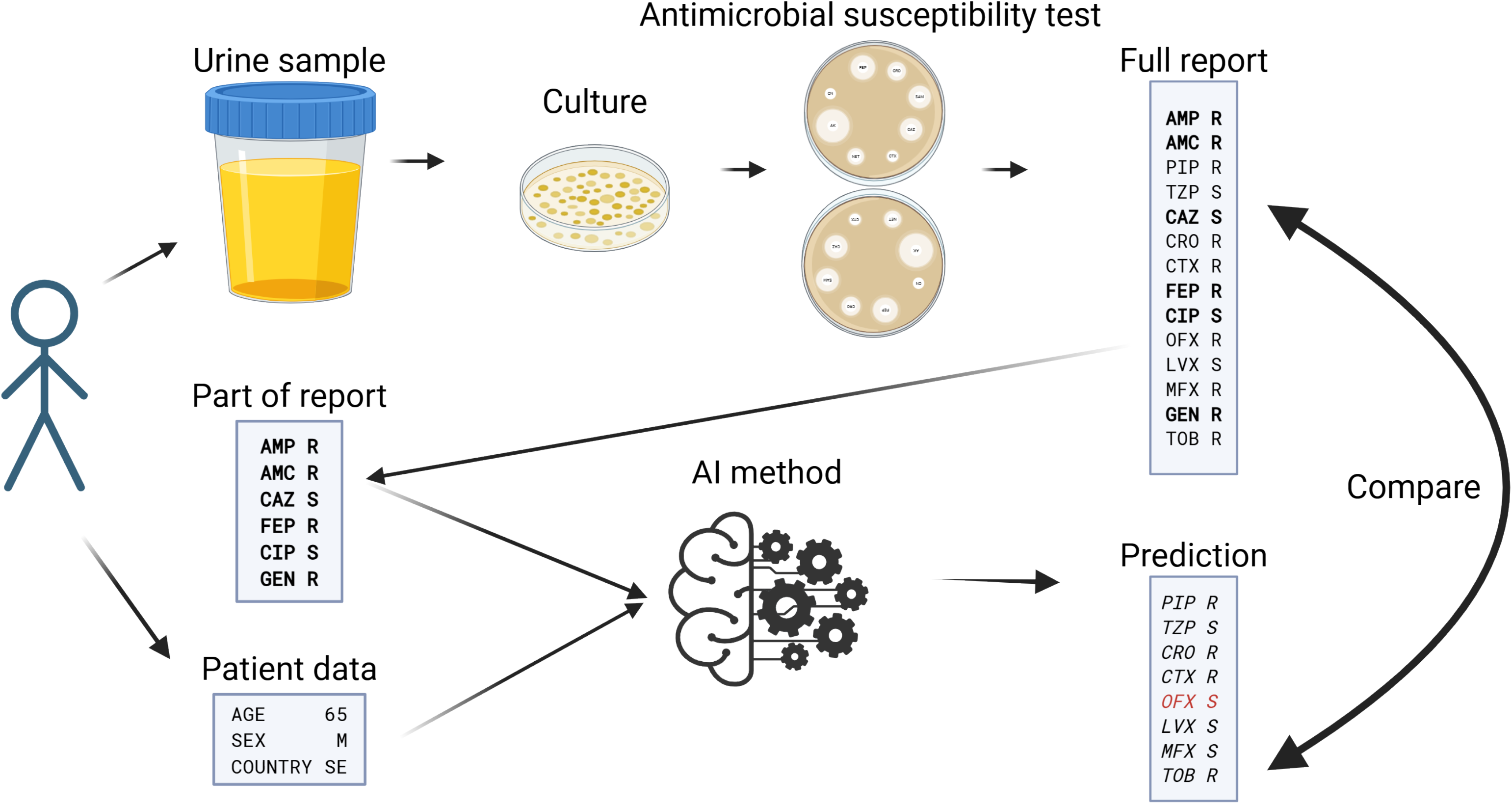
Overview of the clinical evaluation of the AI-based susceptibility prediction method. Urine samples from patients with Escherichia coli bacteriuria were cultured and tested using EUCAST disk diffusion methodology. Susceptibility results for 14 antibiotics were used for evaluation of the AI method. Complete susceptibility profiles served as reference results. For the evaluation of the AI method, subsets of susceptibility test results for 4–8 antibiotics together with patient metadata (age, sex, country, and sampling date) were used as input to predict susceptibility to the remaining antibiotics. Predicted susceptibility categories were compared with the corresponding measured results. Performance estimates were based on all possible combinations in which AST results for 4–8 antibiotics were used as input. **Created in BioRender. Hessel, M.** (**2026**) https://BioRender.com/n1g7fm2. **Licensed under CC BY 4.0.**

### Sample selection process

A structured sample selection process was designed to obtain 100 clinical isolates with diversity in antibiotic susceptibility profiles, patient sex and age. Based on urine culture prevalences at the Department of Clinical Microbiology, Sahlgrenska University Hospital, December 2021–January 2022, weighting factors were assigned for sex, age group, and preliminary susceptibility category to achieve the intended study population. The first isolate of *E. coli* from each eligible patient was included if it met the inclusion criteria (Supplementary Table S1).

### Culture and species identification

*E. coli* isolates included in the evaluation were obtained from urine samples cultured between September 2022 and June 2023. Two agar plates were used, both prepared in-house (Substrate Unit, Department of Clinical Microbiology, Sahlgrenska University Hospital). Species identification and bacterial enumeration were assessed on the first agar plate, which was divided into two half-circle shaped agars, one consisting of chromogenic agar (UriSelect 4, Bio-Rad, Hercules, CA, USA), and the other of horse blood agar supplemented with colistin and nalidixic acid. The second agar plate consisted of Mueller-Hinton agar used for direct susceptibility testing of oral antibiotics (see below). Inoculation was performed by spreading 10 μL of urine on each agar plate using the Kiestra InoqulA+ robot (Becton, Dickinson and Company (BD), Franklin Lakes, NJ, USA). The plates were incubated in a Kiestra Read A Standalone incubator (from BD) at 36°C, for 16–20 hours, the Mueller-Hinton plate in an air environment, and the UriSelect/horse blood plate in air supplemented with 5% CO_2_. All plates were read on a computer screen with Synapsys software from BD. Initial species identification was based on typical pink color of colonies on the chromogenic agar and susceptibility to cefadroxil, or by using VITEK MS MALDI-TOF (bioMérieux, Marcy l’Etoile, France).

#### Antibiotic susceptibility assessment for use in the inclusion algorithm

Initial antibiotic susceptibility testing was performed on plates directly inoculated with urine. Before incubation, the following six disks were applied onto the plate: amoxicillin/clavulanic acid 20/10 μg, cefadroxil 30 μg, ciprofloxacin 5 μg, mecillinam 10 μg, nitrofurantoin 100 μg and trimethoprim 5 μg (all from OXOID, Thermo Fisher Scientific, Waltham, MA, USA). Zone diameters were read digitally and interpreted using EUCAST version 12.0 (2022) breakpoints (4, 18). These preliminary results guided isolate selection but were not used for evaluating the AI method.

#### Antibiotic susceptibility determined by disk diffusion test

Extended AST was performed using the EUCAST disk diffusion methodology, with the reading guideline 9.0 (19) and the breakpoints according to version 13.0, published in 2023 (4). Breakpoints for systemic infection with intravenous administration were used when available. Zone diameter limits used for susceptibility categorization are listed in Supplementary Table S2. Quality control (QC) strains *E. coli* CCUG 17620 and CCUG 30600 were included and compared against the EUCAST QC tables, version 13.0, published in 2023 (4).

Twenty-one antibiotic disks were applied to Mueller-Hinton agar plates. In addition to the six disks also included in the initial susceptibility testing, 15 additional disks were applied: ampicillin 10 μg, cefotaxime 5 μg, ceftazidime 10 μg, ceftriaxone 30 μg, cefepime 30 μg, gentamicin 10 μg, levofloxacin 5 μg, meropenem 10 μg, moxifloxacin 5 μg, nalidixic acid 30 μg, ofloxacin 5 μg, piperacillin 30 μg, piperacillin/tazobactam 30 μg/6 μg, tobramycin 10 μg, and trimethoprim/sulfamethoxazole 1.25 μg/23.75 μg (all from OXOID, Thermo Fisher Scientific). Plates were incubated aerobically at 36°C, for 16–20 h. Inhibition zones were measured digitally using Synapsys software (BD). After antibiotic susceptibility testing was completed, isolates were stored in 10% glycerol at −20°C until further processing.

Among the 21 antibiotics for which inhibition zones were measured, seven were not used in the AI method. Nalidixic acid was used solely for quality control, whereas nitrofurantoin, mecillinam, trimethoprim, cefadroxil, trimethoprim/sulfamethoxazole, and meropenem were used to characterize isolate phenotypes (Fig. 2).

**Figure 2.**
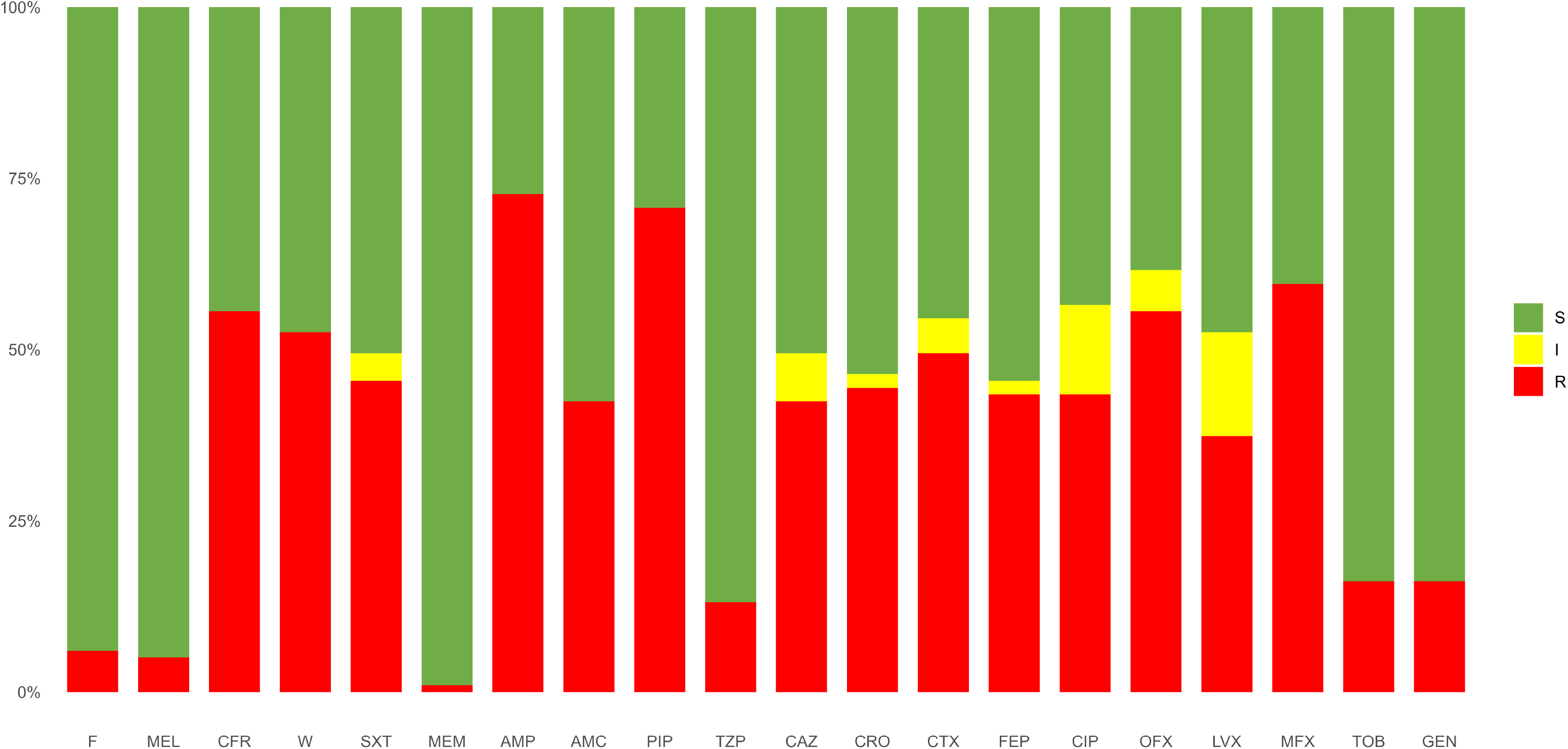
Distribution of EUCAST susceptibility categories among the 99 included E. coli isolates. Bars show the proportions of isolates categorized as susceptible (S), susceptible with increased exposure (I), or resistant (R) for each antibiotic according to EUCAST version 13.0 breakpoints. F, nitrofurantoin; MEL, mecillinam; CFR, cefadroxil; W, trimethoprim; SXT, trimethoprim/sulfamethoxazole; MEM, meropenem; AMP, ampicillin; AMC, amoxicillin/clavulanic acid; PIP, piperacillin; TZP, piperacillin/tazobactam; CAZ, ceftazidime; CRO, ceftriaxone; CTX, cefotaxime; FEP, cefepime; CIP, ciprofloxacin; OFX, ofloxacin; LVX, levofloxacin; MFX, moxifloxacin; GEN, gentamicin; TOB, tobramycin.

### ESBL testing

ESBL classification followed the definitions published by the Public Health Agency of Sweden (20), reflecting the routine diagnostic workflow used in Swedish clinical microbiology laboratories. According to this classification system, isolates resistant to cefotaxime, ceftazidime, or a carbapenem are further investigated for ESBL production. In this classification system, ESBL_A_ denotes clavulanate-inhibited extended-spectrum beta-lactamases, ESBL_M_ denotes plasmid-mediated AmpC beta-lactamases, and ESBL_CARBA_ denotes carbapenemase-producing isolates.

Double disk synergy test for the detection of ESBL was performed on Mueller-Hinton agar plates for all included *E. coli* isolates, but only isolates resistant to cefotaxime, ceftazidime, or meropenem were assessed for ESBL-production. For the detection of ESBL_A_, a disk of amoxicillin/clavulanic acid (20 μg/10 μg) was placed in the center of the agar surrounded by the cefotaxime (30 μg), ceftazidime (30 μg) and cefepime (30 μg) disks (all from OXOID, Thermo Fisher Scientific). Antibiotic Neo-sensitabs disks (Rosco Diagnostica, Taastrup, Denmark) were used for the detection of ESBL_M_ with cloxacillin 500 μg applied between the ceftazidime 30 μg and cefoxitin 30 μg disks. The ESBL_A_ and ESBL_M_ tests were considered positive if there was synergy between any of the applied disks and amoxicillin/clavulanic acid (ESBL_A_) or cloxacillin (ESBL_M_), respectively.

None of the synergy tests used can determine whether a detected ESBL enzyme is encoded by chromosomally located genes or carried by plasmids.

### Heatmaps, clustering, and genotype-error analyses

Heatmaps and hierarchical clustering were used to visualize phenotypic susceptibility patterns based on inhibition zone diameters. Euclidean distance and Ward’s minimum variance method were used for hierarchical clustering of isolates (21). The optimal number of clusters was estimated using the silhouette method (22). Exploratory analyses were performed to investigate associations between resistance determinant groups and isolate-level mean absolute prediction error. Differences in mean absolute prediction error between isolates carrying and lacking each resistance determinant group were assessed using Welch’s t-test, with false discovery rate correction for multiple comparisons. Details of clustering, heatmap generation, and genotype-error analyses are provided in the Supplemental Material.

### Whole-genome sequencing and genomic characterization

Whole-genome sequencing was performed to confirm species identity and to investigate genomic mechanisms associated with AI prediction performance. Genomic DNA was extracted using a modified version of the Marmur protocol (23, 24). Before whole-genome sequencing, isolates were screened by partial 16S rRNA gene Sanger sequencing (25) to detect contaminated DNA preparations and avoid unnecessary sequencing of mixed cultures. Only isolates passing this screening underwent whole-genome sequencing. Sequencing data were subsequently analysed to confirm species identity, assign sequence types using the Achtman *Escherichia coli* seven-gene MLST scheme (26) and identify antimicrobial resistance genes and resistance-associated mutations. Beta-lactamase genes were further classified into functional groups. Detailed laboratory protocols, sequencing procedures, software versions, databases, and bioinformatic parameters are provided in the corresponding sections of the Supplementary Methods.

### AI-based susceptibility prediction

The AI method, developed at Chalmers University of Technology in collaboration with the Fraunhofer-Chalmers Centre, both in Gothenburg, Sweden, was trained on reported antibiotic susceptibility of bloodstream isolates to the European Surveillance System (TESSy) (27), including 1,161,000 *E. coli* (17, 27). Applied in this study, the AI method predicted susceptibility (S) or resistance (R) of an isolate to 14 different antibiotics, namely ampicillin, amoxicillin/clavulanic acid, piperacillin, piperacillin/tazobactam, ceftazidime, ceftriaxone, cefotaxime, cefepime, ciprofloxacin, ofloxacin, levofloxacin, moxifloxacin, gentamicin, and tobramycin, based on known susceptibility data for a subset of antibiotics. Input variables included age, sex, country, sampling date and dichotomized susceptibility test results (S/R) for a subset of antibiotics. The sampling date was set to December 2020, as the AI method could not extrapolate beyond the time span of the training data. Conformal prediction (28) was applied to estimate prediction uncertainty. This enabled the AI method to abstain from predictions when the uncertainty was too high according to predefined confidence thresholds (90%, 95%, and 97.5%).

To enable comparison between EUCAST (4) and AI-based classifications, the three EUCAST categories (susceptible, susceptible with increased exposure, and resistant) were translated into the two categories susceptible (S) and resistant (R). In the primary translation, referred to as Mode A, the EUCAST category “I” was treated as susceptible (S). Two exploratory modes were also defined. In Mode B, “I” was treated as resistant (R), and in Mode C, both “I” and all measurements within the EUCAST area of technical uncertainty (ATU) were treated as resistant (R). Mode A was used as the primary analysis because it follows the EUCAST interpretation of category I as susceptible with increased exposure. Modes B and C were included as additional analyses to assess how alternative translations of EUCAST categories affected prediction performance. Detailed breakpoint mappings for all antibiotics are provided in Supplementary Table S2. Performance was evaluated by comparing predicted susceptibility with measured AST results. Metrics included major error (ME; false-resistant prediction) rate, very major error (VME; false-susceptible prediction) rate, Matthews correlation coefficient (MCC), and F1 score.

## Results

### Study collection and resistance diversity

Out of 125 isolates registered for inclusion, seven were excluded due to repeated samples from already included patients, six were lost, and thirteen were contaminated with other bacteria, including one isolate excluded after whole-genome sequencing. Hence, 99 isolates of *E. coli* cultured from 99 unique patients (female 62%; median age 49, range 0–89 years) were included in the analysis. When compared with the estimated distribution of patients and isolates, more patients aged 18-64 years than expected were included (51% versus 34%). Also, twice as many ESBL-producing isolates (53% vs. 26%) and fewer cefadroxil-susceptible isolates (44% vs. 71%) than expected were included (Supplementary Fig. S1).

The included isolates were diverse in susceptibility to most of the tested antibiotics (Fig. 2). One exception was meropenem, to which 98 (99%) were susceptible (Fig. 2).

In total, 36 (36%) isolates were found to be in the ATU range for one or more antibiotics. For amoxicillin/clavulanic acid, 25 (25%) of the isolates were within ATU range (19 or 20 mm), indicating susceptibility when used intravenously or orally at higher doses for infections originating from the urinary tract. For ciprofloxacin, 13 (13%) of the isolates were within the ATU range, all having zone diameters spanning the whole I category. For piperacillin/tazobactam, 2 (2%) were within the ATU range, all within zone diameters that indicate resistance (R).

Cluster analysis of the included *E. coli* isolates resulted in six phenotypic clusters based on inhibition zone diameters from disk diffusion testing (Fig. 3A). The same clusters were visualized using inhibition zone diameters in a heatmap (Fig. 3B) and EUCAST susceptibility categories in a corresponding category heatmap (Fig. 3C).

**Figure 3.**
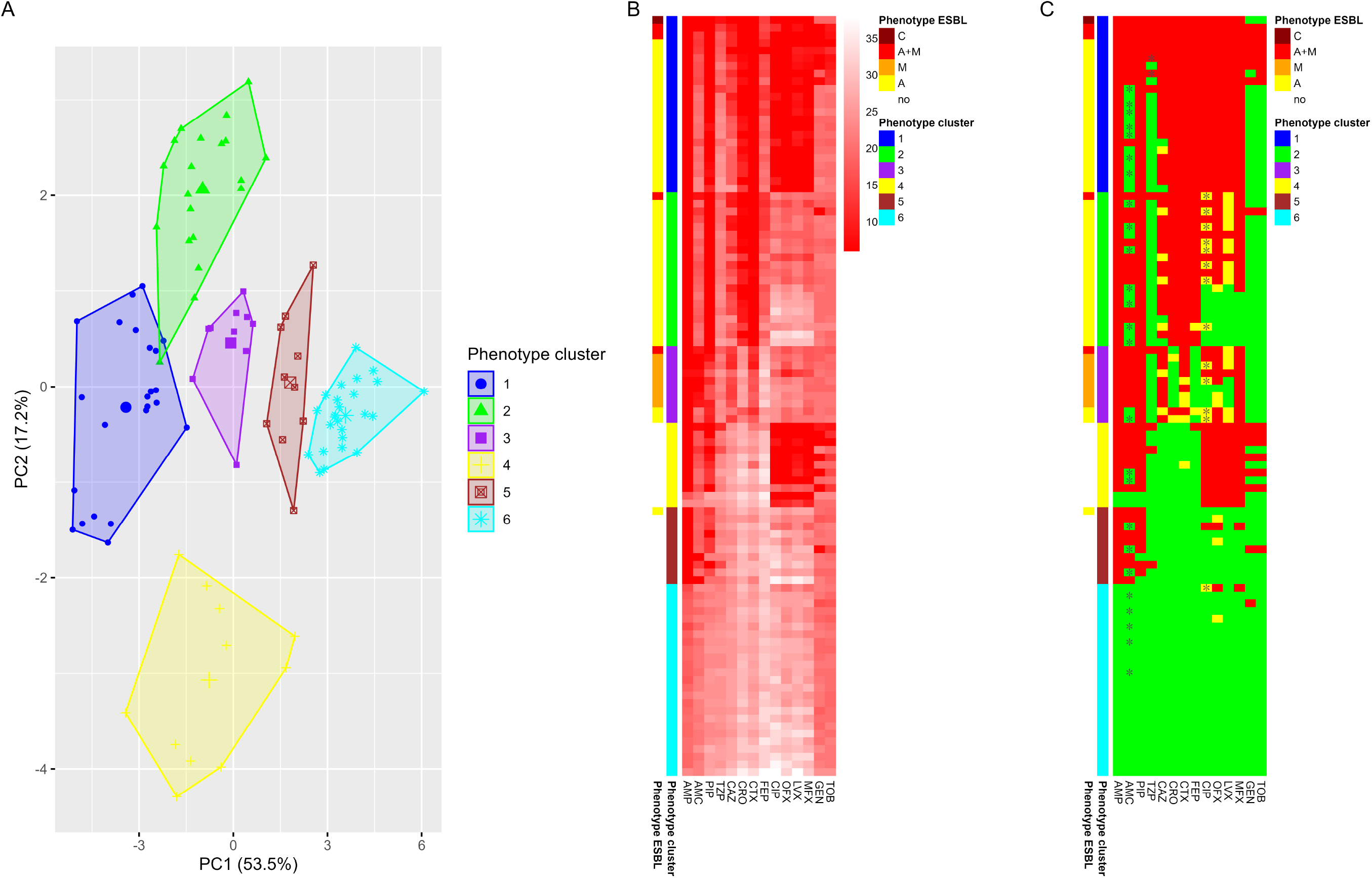
Phenotypic diversity and clustering of the included isolates based on measured inhibition zone diameters. (A) Principal component analysis of inhibition zone diameters showing six phenotype clusters identified by hierarchical clustering. (B) Heatmap of inhibition zone diameters used for clustering. Rows represent isolates and columns represent antibiotics. Darker red colors indicate smaller inhibition zones and thus reduced susceptibility. (C) Corresponding heatmap of EUCAST susceptibility categories for the same isolates and in the same row order as in panel B. Red indicates resistant (R), yellow susceptible with increased exposure (I), and green susceptible (S). Asterisks indicate measurements within the area of technical uncertainty (ATU). Side annotations indicate phenotype cluster assignment and phenotypic ESBL classification. Phenotype cluster: isolate clusters identified from similarities in inhibition zone diameters; Phenotype ESBL: ESBL classification as determined by phenotypic synergy testing; GEN, gentamicin; TOB, tobramycin; AMC, amoxicillin/clavulanic acid; TZP, piperacillin/tazobactam; OFX, ofloxacin; CIP, ciprofloxacin; MFX, moxifloxacin; LVX, levofloxacin; AMP, ampicillin; PIP, piperacillin; CRO, ceftriaxone; CTX, cefotaxime; CAZ, ceftazidime; FEP, cefepime.

### AI susceptibility prediction performance

Evaluation of the AI method was primarily performed in translation mode A, in which the EUCAST category “I” was treated as susceptible (S). Unless otherwise stated, performance was evaluated using AST results for six antibiotics as input to predict susceptibility to the remaining antibiotics.

Without conformal prediction, 80%, 84%, and 86% of predictions were correct when all possible combinations of AST results for four, six, and eight antibiotics, respectively, were evaluated as input (Fig. 4).

**Figure 4.**
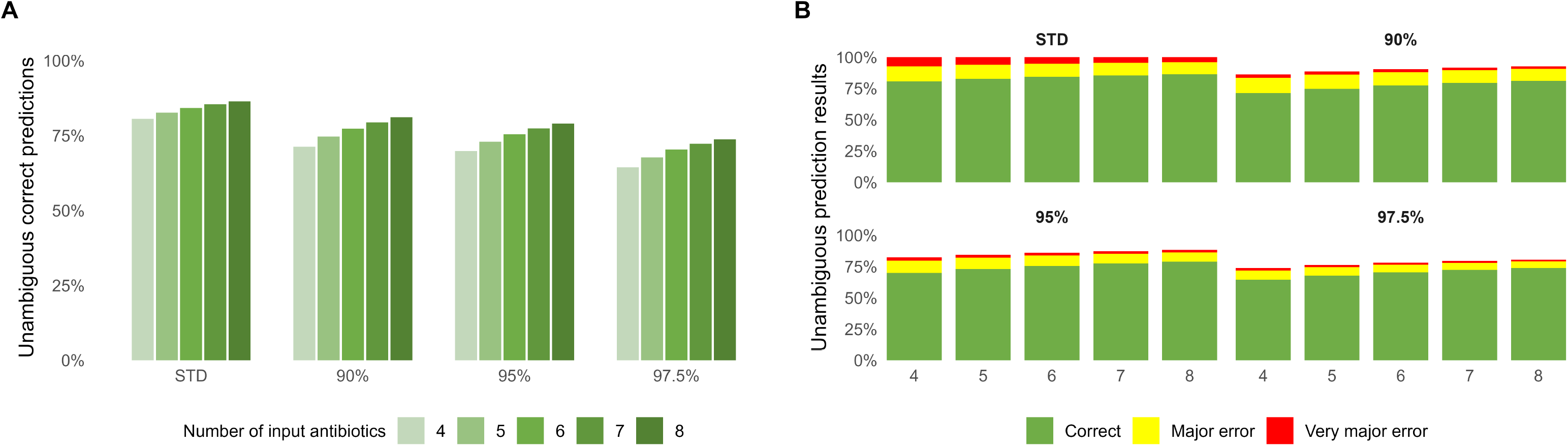
Overall performance of the AI method with and without conformal prediction. (A) Percentage of correct unambiguous predictions obtained using AST results for 4-8 antibiotics as input. STD denotes standard (non-conformal) prediction. Conformal prediction was evaluated at confidence levels of 90%, 95%, and 97.5%, and only predictions assigned a single susceptibility category (S or R) were considered unambiguous. (B) Distribution of prediction outcomes. Green indicates correct unambiguous predictions, yellow major errors (false-resistant predictions), and red very major errors (false-susceptible predictions). Percentages are shown relative to all candidate predictions, including cases where the model abstained from prediction because the required confidence level was not achieved. VME, very major errors; ME, major errors.

Across all predictions generated using AST results for six antibiotics as input, the major error rate (false-resistant predictions) was 19% and the very major error rate (false-susceptible predictions) was 12%. For the included antibiotics, the major error rates ranged from 3% (moxifloxacin) to 51% (amoxicillin/clavulanic acid), and the very major error rates ranged from 3% (levofloxacin) to 26% (piperacillin/tazobactam). Interestingly, prediction of piperacillin showed the highest performance while piperacillin/tazobactam showed the lowest, both according to the Matthews correlation coefficient and F1 score (Fig. 5). At the antibiotic-class level, F1 scores for penicillins, cephalosporins, fluoroquinolones, and aminoglycosides were 0.84, 0.85, 0.87, and 0.57, respectively, compared with 0.74, 0.91, 0.86, and 0.65 in the reference data (17).

**Figure 5.**
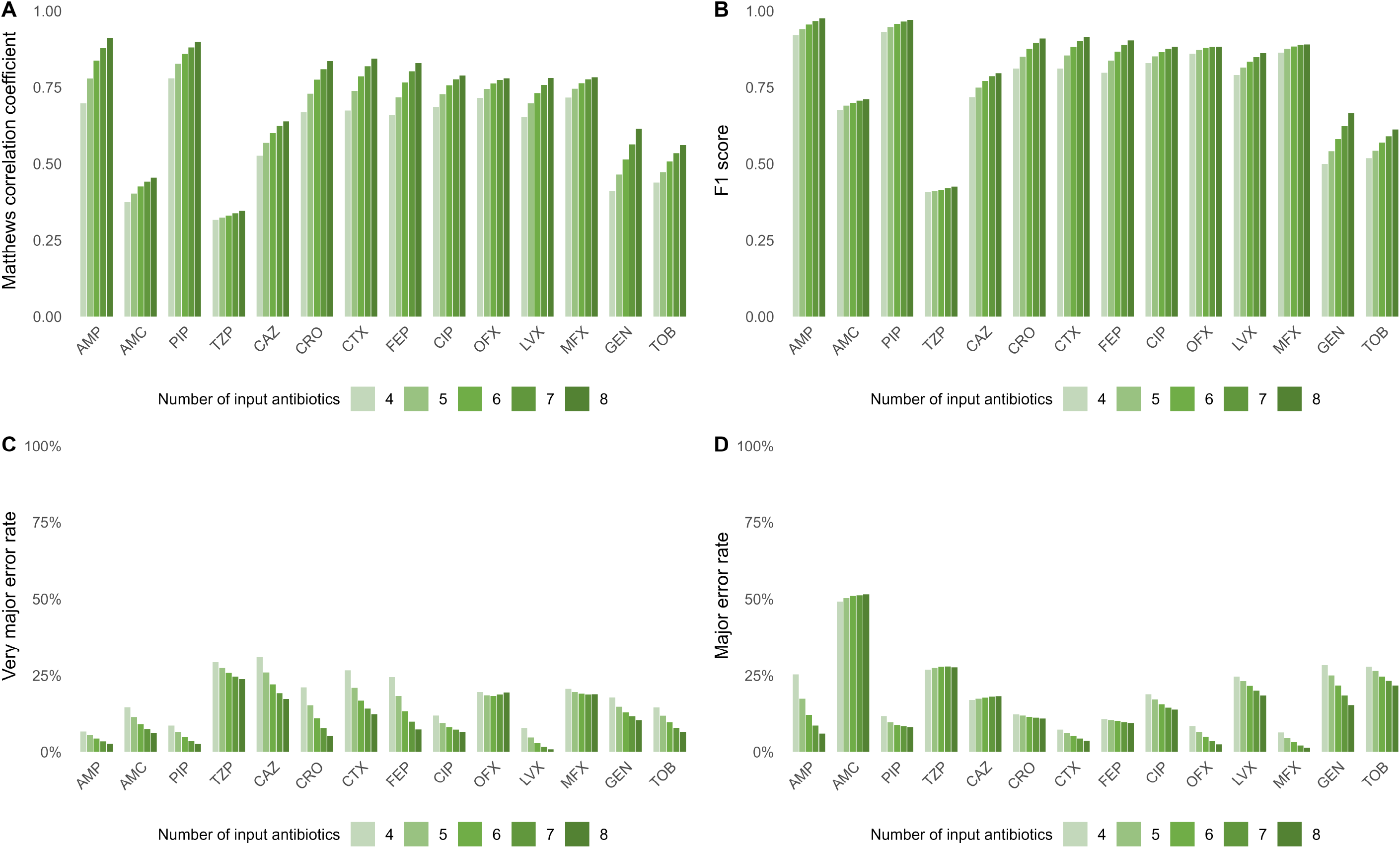
Performance of non-conformal AI predictions for individual antibiotics using four to eight AST results as input. Performance is shown using (A) Matthews correlation coefficient (MCC), (B) F1 score, (C) very major error (VME) rate, and (D) major error (ME) rate. Bars represent prediction performance when 4–8 antibiotics were available as input. Higher MCC and F1 scores indicate better predictive performance, whereas lower VME and ME rates indicate fewer clinically relevant prediction errors. AMP, ampicillin; AMC, amoxicillin/clavulanic acid; PIP, piperacillin; TZP, piperacillin/tazobactam; CAZ, ceftazidime; CRO, ceftriaxone; CTX, cefotaxime; FEP, cefepime; CIP, ciprofloxacin; OFX, ofloxacin; LVX, levofloxacin; MFX, moxifloxacin; GEN, gentamicin; TOB, tobramycin.

When conformal predictions were applied – allowing “S”, “R”, or “Unable to predict with sufficient confidence” as outcomes – the fractions of unambiguous correct predictions, major errors and very major errors decreased as the confidence threshold tightened. At confidence levels of 90%, 95%, and 97.5%, the major error rates were 25%, 16%, and 11%, respectively, while the very major error rates were 10%, 6%, and 4%, respectively. Antibiotic-specific effects of conformal prediction are shown in Supplementary Fig. S3. The method abstained from making predictions in 9.6%, 14%, and 22% of instances at the 90%, 95%, and 97.5% confidence levels, respectively. When the method was applied to the TESSy testing data, the error rates closely matched the confidence levels: the major error rates were 10.1% and 5.1%, and the very major error rates were 9.7% and 5% for the confidence levels of 90% and 95%, respectively. Supplementary Fig. S4 shows details about error rates, F1 and MCC scores for different confidence levels.

The results presented so far were based on all possible combinations in which AST results for 4–8 antibiotics were used as input. Because prediction performance may depend not only on the number but also on the identity of antibiotics used as input, we next restricted the evaluated six-antibiotic combinations to those including at least one antibiotic from each of the four classes: penicillins, cephalosporins, aminoglycosides, and fluoroquinolones. Without conformal prediction, this restriction decreased the major error rate from 19% to 15% and the very major error rate from 12% to 9.5%, while the Matthews correlation coefficient increased from 0.69 to 0.75, with higher values indicating better agreement between predicted and measured susceptibility categories (Supplementary Fig. S7).

Among all evaluated six-antibiotic input combinations, the highest MCC was 0.87. This post hoc best-performing set comprised amoxicillin/clavulanic acid, piperacillin/tazobactam, cefotaxime, ciprofloxacin, ofloxacin, and tobramycin. Using AST results for these six antibiotics as input to predict susceptibility to the remaining antibiotics resulted in a major error rate of 8.3% and a very major error rate of 4.4% (Supplementary Fig. S8). A summary of the different selections of AST results for six antibiotics is shown in Table 1.

**Table 1.** Performance of the AI-based susceptibility prediction method using different six- antibiotic AST input strategies.

| AST input strategy | No. of input combinations | Predicted antibiotics per combination | MCC | ME rate | VME rate |
| --- | --- | --- | --- | --- | --- |
| All six-antibiotic AST input combinations | 3,003 | 8 | 0.69 | 19% | 12% |
| Class-constrained six-antibiotic AST input combinations | 1,536 | 8 | 0.75 | 15% | 9.5% |
| Fixed optimized six-antibiotic AST input set | 1 | 8 | 0.87 | 8.3% | 4.4% |

AST results for six of 14 antibiotics were used as input to predict susceptibility to the remaining eight antibiotics. Class-constrained input combinations required at least one antibiotic from each of four antibiotic classes: penicillins, cephalosporins, fluoroquinolones, and aminoglycosides. The post hoc best-performing input set was defined as the six-antibiotic combination with the highest MCC in this evaluation of 99 isolates; it should not be interpreted as an input set optimized by the AI model or as generally optimal beyond this dataset. This input set consisted of amoxicillin/clavulanic acid, piperacillin/tazobactam, cefotaxime, ciprofloxacin, ofloxacin, and tobramycin. AST, antibiotic susceptibility testing; MCC, Matthews correlation coefficient; ME, major error; VME, very major error.

The AI method was also evaluated in two additional translation modes. In Mode B, the EUCAST category “I” was treated as resistant (R). In Mode C, both “I” and measurements within the ATU range were treated as resistant (R) (Supplementary Table S2). Overall, the major error rates were 20% in both Mode B and C, compared with 19% in Mode A, and the very major error rates were 8% in both Mode B and C, compared with 12% in Mode A. The MCC score was 0.72 in both Mode B and C, compared with 0.69 in Mode A. This pattern is consistent with a more conservative translation of EUCAST categories, where fewer false-susceptible predictions are obtained at the cost of slightly more false-resistant predictions. Antibiotic-specific differences between translation modes were more pronounced than the overall differences (Supplementary Fig. S9).

### Characterization of isolates associated with prediction errors

To further explore the incorrect predictions, a heatmap showing error frequencies was compared with a heatmap of the susceptibility phenotypes. Higher color intensity indicates a higher frequency of incorrect predictions for a given isolate–antibiotic pair across all evaluated six-antibiotic input combinations (Fig. 6A). As expected, the prediction errors followed the patterns within the antibiotic classes cephalosporins, fluoroquinolones, and aminoglycosides. For penicillins and penicillin/beta-lactamase inhibitor combinations, both the type of penicillin and the presence of a beta-lactamase inhibitor affected the error rates. Some isolates susceptible to all antibiotics (Fig. 6A, Cluster 6) showed a few major errors for piperacillin and, to a larger extent, for ampicillin. Conversely, most isolates resistant to all antibiotics (Fig. 6A, part of Cluster 1) showed some very major errors for piperacillin/tazobactam. Consequently, regardless of the measured susceptibility category, some six-antibiotic input combinations always predicted ampicillin, piperacillin, or both as resistant, whereas others always predicted piperacillin/tazobactam as susceptible.

**Figure 6.**
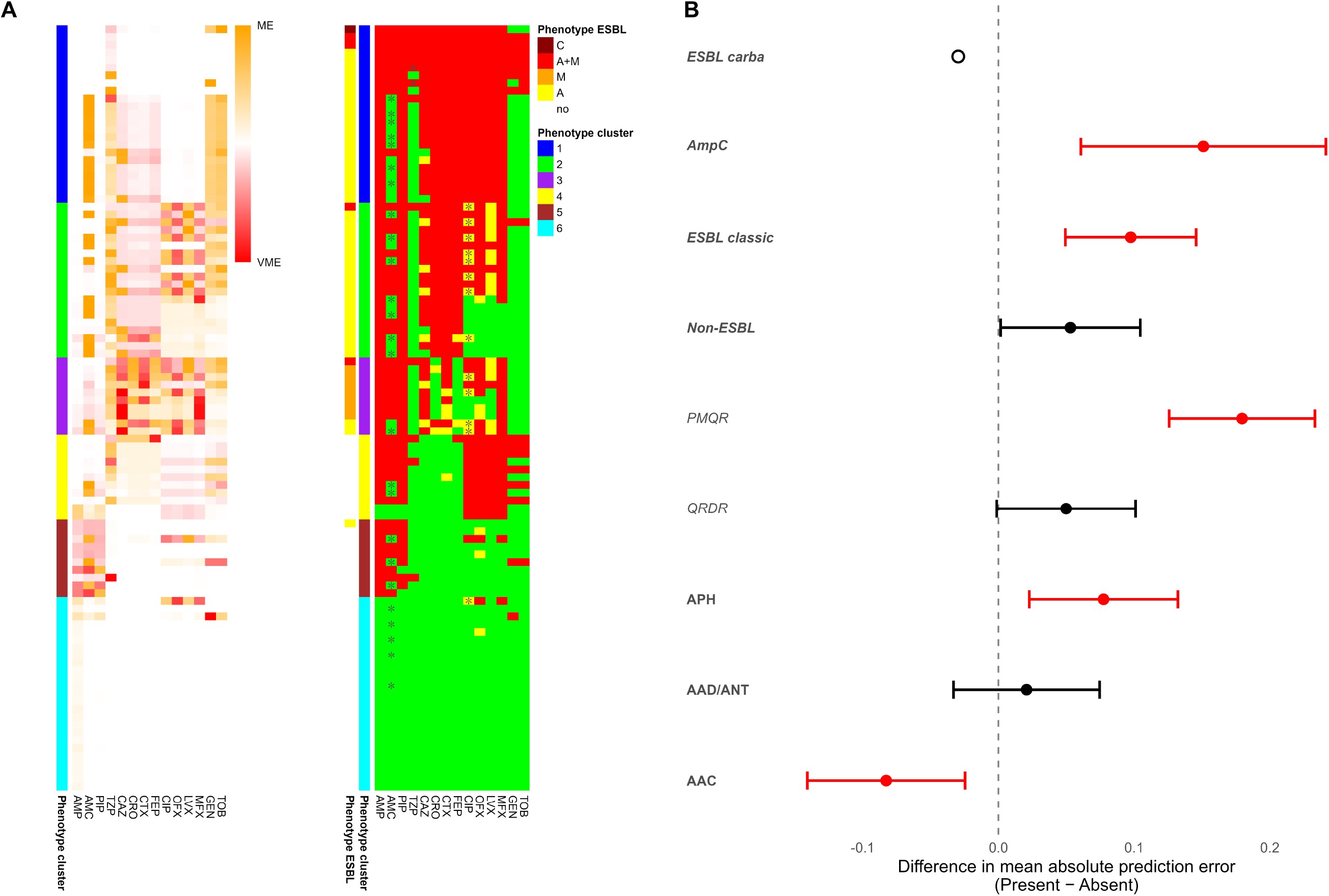
Prediction errors across isolates and associations with resistance determinants. (A) Distribution of major and very major errors across isolates and antibiotics using AST results for six antibiotics as input. Rows represent individual isolates and columns represent antibiotics. The left heatmap shows the frequency of prediction errors across all evaluated combinations in which AST results for six antibiotics were used as input. Orange indicates major errors (false-resistant predictions), and red indicates very major errors (false-susceptible predictions). The right panel provides phenotypic annotations to facilitate interpretation of error patterns, including phenotype cluster assignment derived from hierarchical clustering of inhibition zone diameters, phenotypic ESBL classification, and EUCAST susceptibility categories. Isolates are ordered according to hierarchical clustering based on inhibition zone diameters. (B) Differences in mean absolute prediction error associated with resistance determinant groups. Points represent differences in isolate-level mean absolute prediction error between isolates carrying and lacking a given resistance determinant group (Present − Absent). Positive values indicate higher prediction error among carriers of the resistance determinant, whereas negative values indicate lower prediction error among carriers. Error bars indicate 95% confidence intervals. Mean absolute prediction error was calculated across all antibiotics and evaluated combinations of AST results for six antibiotics as input. Red error bars indicate statistically significant differences according to Welch’s t-test after false discovery rate correction. AMP, ampicillin; AMC, amoxicillin/clavulanic acid; PIP, piperacillin; TZP, piperacillin/tazobactam; CAZ, ceftazidime; CRO, ceftriaxone; CTX, cefotaxime; FEP, cefepime; CIP, ciprofloxacin; OFX, ofloxacin; LVX, levofloxacin; MFX, moxifloxacin; GEN, gentamicin; TOB, tobramycin; VME, very major errors; ME, major errors; QRDR, quinolone resistance-determining region mutations; PMQR, plasmid-mediated quinolone resistance genes; AAC, aminoglycoside N-acetyltransferases; APH, aminoglycoside O-phosphotransferases; AAD, aminoglycoside adenyltransferases; ANT, aminoglycoside nucleotidyltransferases; ESBL carba, carbapenemases; AmpC, AmpC beta-lactamases; ESBL classic, extended-spectrum beta-lactamases excluding AmpC beta-lactamases and carbapenemases; Non-ESBL, beta-lactamases without extended-spectrum activity.

By ranking isolates according to their mean absolute prediction error, calculated as the mean of major and very major error rates across antibiotics, phenotypic patterns associated with predictive performance could be visualized (Supplementary Fig. S12A). Resistance determinant groups were used to assess genotype-associated differences in prediction performance (Supplementary Fig. S12B). Isolates carrying AmpC genes showed higher mean absolute prediction errors than non-carriers (29% versus 14%). Similarly, isolates carrying one or more plasmid-mediated quinolone resistance (PMQR) genes had higher mean absolute prediction errors than isolates lacking such genes (30% versus 12%). In contrast, isolates carrying aminoglycoside N-acetyltransferase (AAC) genes showed lower mean absolute prediction errors than isolates lacking AAC genes (9% versus 17%) (Fig. 6B).

Higher mean absolute prediction errors were observed in phenotype Cluster 2 and 3 (Fig. 6A), corresponding primarily to ESBL-like and AmpC-like phenotypes, respectively. Cluster 2 was characterized by variable low-level fluoroquinolone resistance, while Cluster 3 showed variable cephalosporin susceptibility together with variable low-level fluoroquinolone resistance (Fig. 3). In the genotypic analysis, higher error rates were also observed among isolates carrying AmpC beta-lactamase genes or plasmid-mediated quinolone resistance genes.

## Discussion

Overall, the AI method demonstrated promising predictive performance, comparable to reference data, in this prospective clinical evaluation including 99 urine isolates of *E. coli* representing a broad spectrum of patients and antibiotic resistance profiles. Prediction failures were concentrated to specific phenotypic and genotypic resistance patterns, providing insight into situations where the model was more likely to perform poorly.

The AI method was more prone to perform major errors, corresponding to false-resistant predictions, than to very major errors, corresponding to false-susceptible predictions. As expected, the performance improved with an increased number of AST results used as input to the model. The use of conformal predictions further reduced error rates by restricting predictions to cases with sufficiently low uncertainty, resulting in more conservative but, in general, more reliable results.

In conformal prediction, the expected error rate is 100% minus the predefined confidence level. However, in our evaluation, errors were unevenly distributed, with major errors occurring more often than very major errors, indicating a general tendency of the AI method to predict resistance both when confidence thresholds were used and not used. F1 scores were also slightly higher in the reference data than in our clinical evaluation. This likely reflects differences in patient populations between the data used to train the method and our evaluation data. Also, the training data was based on invasive bloodstream infections, while the isolates in our evaluation derived from urine samples. Indeed, as with language models, predictions reflect patterns in the training data and may behave differently when applied to clinical data with other characteristics.

Sample-level heatmaps gave further insight into the performance of the AI method. For some isolates with all-susceptible phenotypes, certain six-antibiotic input combinations resulted in resistant predictions for piperacillin and ampicillin. Conversely, for some isolates with all-resistant phenotypes, certain input combinations resulted in susceptible predictions for piperacillin/tazobactam. In a clinical application, such input combinations should be avoided. This study was not designed to identify the optimal set of six AST results to use as input, but when we restricted the input set to combinations with at least one antibiotic per class, the performance measure MCC increased from 0.69 to 0.75. When all six-antibiotic input combinations were ranked by MCC, the highest MCC was 0.87, suggesting that optimizing the antibiotics included in the initial susceptibility testing panel could improve performance in a clinical setting.

The higher error rates in the ESBL-like and AmpC-like phenotype clusters may reflect the difficulty of predicting intermediate-complexity resistance profiles from partial phenotypic input. These isolates were neither uniformly susceptible nor broadly resistant across the full antibiotic panel but instead showed variable susceptibility within antibiotic classes. Since the AI method relies on correlations between susceptibility patterns, this within-class variability may impair prediction of susceptibility to other antibiotics within the same class.

Susceptibility testing of amoxicillin/clavulanic acid is known to be problematic for several reasons. First, breakpoints differ between CLSI and EUCAST (29, 30). Second, EUCAST applies different breakpoints and interpretations for different indications and route of antibiotic administration (4). These factors may have influenced the quality and consistency of the TESSy data used for training of the AI model, particularly for amoxicillin/clavulanic acid.

To our knowledge, no prior studies have evaluated antibiotic susceptibility prediction based solely on known susceptibility to other antibiotics. Most existing machine learning approaches instead rely on genomic, proteomic, metagenomic, or electronic health record data to predict MICs, categorical susceptibility, or patient-level resistance risk (7–15). These studies illustrate the diversity of data sources used in AI-based antimicrobial resistance prediction. Our study adds a different perspective by evaluating whether susceptibility to some antibiotics can be leveraged to predict susceptibility to others in prospectively collected clinical isolates.

A strength of this study was the structured sample-selection process, designed to increase diversity while reducing subjective selection bias. The resulting high-resistance collection may have provided a clinically challenging dataset for evaluating the robustness of the model. Despite the structured approach, more resistant isolates and patients aged 18–64 years were included than expected, which may have introduced selection bias.

The breakpoints used for susceptibility testing in the TESSy data, used to train the AI model, are unknown. First, it is unclear whether EUCAST guidelines were consistently applied. Second, EUCAST breakpoints have changed over time, and laboratories may adopt such changes with some delay. Each result, however, includes a date and country code, which allows the AI method to potentially learn implicit adjustments for temporal changes and national practices. As a result, training data from other countries may have less influence on predictions than Swedish data. Geographic and temporal effects may reflect both differences in breakpoint usage and variations in circulating strains. Additionally, the EUCAST I category (Susceptible, increased exposure) was not included as a separate prediction category in the AI method. The results from modes B and C show that performance estimates depend on how EUCAST categories are translated to the binary S/R format required by the AI method. More conservative translations, in which category I and ATU measurements were treated as resistant, reduced false-susceptible predictions but slightly increased false-resistant predictions. This should not be interpreted as an intrinsic improvement of the AI method, since it partly reflects a changed definition of susceptibility and resistance. Therefore, mode A was retained as the primary analysis. Another limitation of the AI method is the limited information it uses. For each patient, 4–8 antibiotic susceptibility test results (S/R) were provided as input, along with sex, age, country, and date. While these demographic and temporal variables may reflect prevalence differences, they do not represent biological properties of the isolates and are unlikely to reveal mechanistic links between resistance to different antibiotics, including beta-lactams, fluoroquinolones and aminoglycosides.

Another limitation of the method is its lack of explainability. Neural networks function as ‘black boxes’ and therefore provide limited information about which inputs drive specific predictions. More interpretable approaches, such as gradient boosting decision trees applied to electronic medical records (14) or genome-derived sequence features (11), could provide information about what variables contribute to the predictions. In the latter, the most informative DNA substrings (k-mers) could be linked to beta-lactamase genes. Although neural networks are inherently less transparent, techniques for improving model explainability are available (31).

Future refinements of the evaluated AI model could incorporate additional diagnostic data (e.g., genomic, proteomic, or clinical). Using raw values such as zone diameters or MICs would allow the AI method to predict zone diameters or MICs, independent of current breakpoints. Significant information may be lost when zone diameters are converted into categorical susceptibility test results. Uncertainty control methods analogous to conformal prediction could still be applied to provide confidence intervals for predicted values. As AI technology evolves, alternative model architectures beyond transformers could also be explored.

## Conclusion

Although the diversity of included *E. coli* isolates differed from expectations, the inclusion algorithm proved effective for selecting bacterial isolates with varied antibiotic susceptibility from patients of different ages and sexes. The AI method, previously shown to accurately predict antibiotic susceptibility from historical data, correctly predicted susceptibility for most antibiotics in clinical *E. coli* isolates in a Swedish setting. With further refinement, such as the use of raw zone diameters, prediction of three susceptibility categories (S/I/R) or integration of additional diagnostic data, this approach could become a valuable tool for clinical decision support and antimicrobial stewardship.

## Data Availability

Raw sequencing reads and genome assemblies generated in this study have been deposited in the European Nucleotide Archive (ENA) under BioProject accession number PRJEB115520. Additional isolate-level data supporting the findings are provided in the Supplementary Material.

https://www.ebi.ac.uk/ena/browser/view/PRJEB115520

## Acknowledgements

We thank Lennart Svensson for helpful scientific discussions about machine learning, and the laboratory technician Niklas Eklo for coordinating the inclusion of eligible subjects and performing additional antibiotic susceptibility testing of isolates.

## Ethical statement

The Swedish Ethical Review Authority (no 2022-05318-01) approved the study. No informed consent was required for this study.

## Data Availability

Raw sequencing reads and genome assemblies generated in this study have been deposited in the European Nucleotide Archive (ENA) under BioProject accession number PRJEB115520.

