## Supplemental Material for "Clinical evaluation of artificial intelligence for diagnostics of antibiotic-resistant bacteria"

#### Supplementary Methods

##### Heatmaps, clustering, and exploratory genotype-error analyses

Antibiotic susceptibility patterns were illustrated by heatmaps and clustering. The zone diameter measured for each antibiotic in millimeters was used as the basis for heatmaps and cluster plots. Isolates were shown in rows on heatmaps, and the row order from the initial clustering was kept across all heatmaps.

Euclidean distance and Ward's minimum variance method were used for hierarchical clustering of isolates (1). Hierarchical clustering was performed for both isolates and antibiotics, but only isolate clustering was used for subsequent analyses and figure annotations. The optimal number of clusters was estimated using the silhouette method (2). Clusters were visualized with principal component analysis and heatmaps. Analyses were performed in R, version 4.5.2, using the packages factoextra, version 1.0.7, and pheatmap, version 1.0.13.

To investigate associations between resistance mechanisms and prediction performance, isolates were stratified according to the presence or absence of specific resistance mechanism groups, including AmpC beta-lactamases, plasmid-mediated quinolone resistance genes, aminoglycoside N-acetyltransferases, and quinolone resistance-determining region mutations. Mean absolute prediction error was calculated for each isolate across all antibiotics and evaluated prediction combinations. Differences between groups were assessed using Welch's t-test with false discovery rate correction. These analyses were exploratory.

#### DNA extraction

Included isolates stored in 10% glycerol at −20°C were cultured on one non-selective agar plate (Columbia Blood Agar Base plus 5% defibrinated horse blood, prepared in-house at the Substrate Unit, Department of Clinical Microbiology, Sahlgrenska University Hospital, Gothenburg, Sweden) and one chromogenic agar plate (UriSelect 4, Bio-Rad, Hercules, CA, USA), both incubated overnight at 36°C in air. Both media were used for contamination checks, but the biomass used for DNA extraction was exclusively harvested from the horse blood agar.

Genomic DNA was extracted using a modified version of the Marmur protocol (3, 4). Cells were lysed using 40 µL of Proteinase K (Qiagen, Venlo, Netherlands) with >600 mAU/mL enzymatic activity. DNA quality was assessed using a NanoDrop spectrophotometer (Thermo Fisher Scientific, Waltham, MA, USA), and DNA concentration was quantified using a Qubit fluorometer, with a Broad Range kit (Thermo Fisher Scientific).

#### 16S rRNA gene screening

Prior to whole-genome sequencing, DNA extracts were screened by partial 16S rRNA gene Sanger sequencing (5) to detect contaminated DNA preparations and avoid unnecessary whole-genome sequencing of mixed cultures. PCR amplification of a fragment between positions 28 and 1494 of the *E. coli* 16S rRNA gene was performed using a Bioer GeneTouch PCR machine version 1.1 (Bioer Technology, Hangzhou, Zhejiang, China) with 1 µL of DNA diluted 1:10. The PCR protocol consisted of an initial denaturation step at 95°C for 2 min, followed by 30 cycles of denaturation at 95°C for 30 s, annealing at 55°C for 1 min, and elongation at 72°C for 2 min, and a final elongation step at 72°C for 10 min (6). The primers used were 16F28 (AGAGTTTGATCKTGGCTCAG) and 16r1494 (TACGGYTACCTTGTTACGAC) (7, 8). The PCR product was Sanger sequenced using primer 16R806 (GGACTACCAGGGTATCTAAT)(9).

Species screening was performed using multiBLAST against a 16S rRNA gene database available in CLC Genomics Workbench (Qiagen, Venlo, Netherlands). Results were accepted if the top hit showed ≥99% identity to a species within the genera *Escherichia* or *Shigella* and there was no chromatogram evidence of contamination. Because partial 16S rRNA gene sequencing was used only as a screening step, definitive species identification was based on whole-genome sequencing analyses.

#### Whole-genome sequencing

Extracted DNA that passed the Sanger 16S rRNA screening was sent to Eurofins Genomics (Konstanz, Germany) for preparation of a standard genomic Illumina library. Whole-genome sequencing was performed using an Illumina NovaSeq 6000 platform (Illumina, San Diego, CA, USA), producing between 7 and 30 million paired-end reads with a length of 151 base pairs.

#### Bioinformatic analysis and resistance gene identification

Sequencing reads were screened for contamination using ConFindr, version 0.8.2 (10). The reads were trimmed and quality-checked with Trim Galore, version 2.2.0 (11), which used Cutadapt, version 5.2 (12), and FastQC, version 0.12.1 (13). Trimmed reads were assembled with SPAdes, version 4.2.0 (14). Assembly quality was evaluated using QUAST, version 5.3.0 (15), and further contamination assessment was performed using CheckM, version 1.2.5 (16). Average nucleotide identity based on BLAST (ANIb) with the *E. coli* type strain DSM 30083$^{T}$ was assessed using the web service JSpeciesWS (17), and final species identification was performed using the web service Type Strain Genome Server (TYGS) (18). The latter two web services were accessed in December 2023. MultiQC, version 1.35 (19), was used to summarize sequencing quality metrics from Cutadapt, FastQC, and QUAST. Sequence types were assigned from genome assemblies using mlst version 2.35.0 (https://github.com/tseemann/mlst) with the Achtman *Escherichia coli* seven-gene MLST scheme (adk, fumC, gyrB, icd, mdh, purA, and recA) (20, 21). To ensure use of current allele and profile definitions, the EnteroBase Escherichia.Achtman7GeneMLST allele FASTA files and profile table were downloaded July 13, 2026 and converted into an mlst-compatible local database before analysis (22).

Antimicrobial resistance genes and resistance-associated point mutations were identified using AMRFinderPlus, version 4.2.7 (23), with the curated core AMR gene database release May 15, 2026. In cases where gene identification was incomplete or ambiguous, a complementary analysis was performed using ARIBA, version 2.14.7 (24), with the ResFinder database release May 25, 2026 (25), on quality-filtered sequencing reads. Beta-lactamase genes were further classified into functional groups (26) based on the Beta-Lactamase DataBase (BLDB) (27) accessed June 24, 2026.

### Supplementary Tables

| **Supplementary Table S1 – Inclusion algorithm and sample selection** | | | |
|---|---|---|---|
| **Property** | **Percentage** | **Definition** | **Rule for inclusion** |
| *Patients* | | | |
| Sex: Male | 100% | NA | PID second last digit 1, 3, 5, 7 or 9 |
| Sex: Female | 40% | NA | PID second last digit 0 or 2 |
| Age: 0–17 years | 100% | NA | PID any last digit |
| Age: 18–64 years | 50% | NA | PID last digit 0, 1, 2, 3 or 4 |
| Age: ≥65 years | 30% | NA | PID last digit 0, 1 or 2 |
| *Antibiotic susceptibility of bacterial isolates* | | | |
| Category I | 100% | ESBL-producing isolate resistant to either cefotaxime or ceftazidime | LIS sample number: Any last digit |
| Category II | 100% | Non-ESBL-producing isolate resistant to either cefotaxime or ceftazidime | LIS sample number: Any last digit |
| Category III | 100% | Isolate resistant to cefadroxil, but neither to cefotaxime nor ceftazidime | LIS sample number: Any last digit |
| Category IV | 100% | Isolate susceptible (S) to cefadroxil, and non-S to at least two of ciprofloxacin, nitrofurantoin, trimethoprim or mecillinam | LIS sample number: Any last digit |
| Category V | 10% | Isolate susceptible (S) to cefadroxil, and non-S to at most one of ciprofloxacin, nitrofurantoin, trimethoprim or mecillinam | LIS sample number: Last digit is 0 |

***Inclusion algorithm and sample selection.*** *To obtain a diverse set of Escherichia coli isolates, inclusion rules were applied based on patient sex, age, and preliminary susceptibility profiles. Sex was determined from the second-to-last digit of the Swedish 12-digit patient identification number (PID) (even — female, odd — male), while age distribution was approximated from the last digit, which—together with the second-to-last digit—is evenly distributed in the Swedish population. Additional rules for antibiotic susceptibility were derived from the laboratory information system (LIS) sample number. Isolates were included if they passed preliminary oral antibiotic susceptibility screening and matched the required PID and LIS digit criteria, which were used only to adjust prevalence, except for the second-to-last PID digit indicating sex. Susceptibility categories I to V represent different outcomes in the laboratory process of screening and confirming ESBL-producing isolates from the urinary tract and susceptibility to oral antibiotics. The percentages shown indicate the average fraction of eligible samples* *expected to pass the inclusion rules. NA, Not applicable; PID, Swedish patient identification number; LIS, Laboratory information system; S, susceptible; Non-S, resistant (R) or susceptible increased exposure (I).*

**Supplementary Table S2 – Zone diameter breakpoints applied for the AI method**

| Antibiotic | EUCAST 13.0 | | | | AI method | | | | | |
|---|---|---|---|---|---|---|---|---|---|---|
| | | | | | Mode A | | Mode B | | Mode C | |
| | S | I | R | ATU | S | R | S | R | S | R |
| AMP | ≥14 | | <14 | | ≥14 | <14 | ≥14 | <14 | ≥14 | <14 |
| AMC | ≥19 | | <19 | 19–20 | ≥19 | <19 | ≥19 | <19 | ***≥21*** | ***<21*** |
| PIP | ≥20 | | <20 | | ≥20 | <20 | ≥20 | <20 | ≥20 | <20 |
| TZP | ≥20 | | <20 | 19 | ≥20 | <20 | 20 | <20 | ≥20 | <20 |
| CAZ | ≥22 | 19–21 | <19 | | ≥19 | <19 | ***≥22*** | ***<22*** | ***≥22*** | ***<22*** |
| CRO | ≥25 | 22–24 | <22 | | ≥22 | <22 | ***≥25*** | ***<25*** | ***≥25*** | ***<25*** |
| CTX | ≥20 | 17–19 | <17 | | ≥17 | <17 | ***≥20*** | ***<20*** | ***≥20*** | ***<20*** |
| FEP | ≥27 | 24–26 | <24 | | ≥24 | <24 | ***≥27*** | ***<27*** | ***≥27*** | ***<27*** |
| CIP | ≥25 | 22–24 | <22 | 22–24 | ≥22 | <22 | ***≥25*** | ***<25*** | ***≥25*** | ***<25*** |
| OFX | ≥24 | 22–23 | <22 | | ≥22 | <22 | ***≥24*** | ***<24*** | ***≥24*** | ***<24*** |
| LVX | ≥23 | 19–22 | <19 | | ≥19 | <19 | ***≥23*** | ***<23*** | ***≥23*** | ***<23*** |
| MFX | ≥22 | | <22 | | ≥22 | <22 | ≥22 | <22 | ≥22 | <22 |
| GEN | ≥17 | | <17 | | ≥17 | <17 | ≥17 | <17 | ≥17 | <17 |
| TOB | ≥16 | | <16 | | ≥16 | <16 | ≥16 | <16 | ≥16 | <16 |

*Zone diameter breakpoints (mm) used to categorize isolates into S (susceptible) and R (resistant) for the AI method, based on EUCAST version 13.0 breakpoints. In translation Mode A, EUCAST's S and I categories were grouped as S, while R remained R. In Mode B, EUCAST's S category was treated as S, and both I and R as R. In Mode C, measurements within the area of technical uncertainty (ATU) were also considered, with isolates in I, R, or ATU (within the S range) being treated as R. Breakpoints shown in* ***bold italics*** *in columns Mode B and Mode C indicate differences in translation compared with Mode A. AMP, ampicillin; AMC, amoxicillin/clavulanic acid; PIP, piperacillin; TZP, piperacillin/tazobactam; CAZ, ceftazidime; CRO, ceftriaxone; CTX, cefotaxime; FEP, cefepime; CIP, ciprofloxacin; OFX, ofloxacin; LVX, levofloxacin; MFX, moxifloxacin; GEN, gentamicin; TOB, tobramycin.*

**Supplementary Table S3. Isolate collection dates, accessions, sequence types, and resistance determinants.**

The table is provided as a separate Excel file. It links the isolate identifiers used in the manuscript to the corresponding collection dates and ENA sample, experiment, and run accessions, and summarizes sequence types and detected resistance determinants. Sequence types were assigned using the Achtman *Escherichia coli* MLST scheme. Beta-lactamase functional classes are reported separately from the detected beta-lactamase genes. Quinolone resistance determinants include plasmid-mediated quinolone resistance genes and quinolone resistance-determining region mutations. QRDR mutations are also included in the combined resistance determinant column. Age and sex are not included in this table. Constants such as organism, isolation source, and country are not included because they are identical for all isolates. The first sheet, 'README', provides general information; the second, 'Column descriptions', defines the columns; and the third, 'Data', contains the table, with one row per included isolate.

### Supplementary Figures

***Supplementary Figure S1. Expected and observed distributions of included isolates****. (A) Distribution of age and sex categories among isolates estimated to be included according to the inclusion algorithm and among isolates ultimately included in the study. Expected proportions were estimated from urine culture statistics collected between December 2021 and January 2022 at the Department of Clinical Microbiology, Sahlgrenska University Hospital, and the predefined inclusion* *algorithm. (B) Distribution of susceptibility categories used for isolate selection. Category I, ESBL-producing isolates; Category II, non-ESBL-producing isolates resistant to either cefotaxime or ceftazidime; Category III, isolates resistant to cefadroxil but not resistant to cefotaxime or ceftazidime; Category IV, isolates susceptible to cefadroxil and non-S to at least two of ciprofloxacin, nitrofurantoin, trimethoprim, or mecillinam; Category V, isolates susceptible to cefadroxil and non-S to at most one of ciprofloxacin, nitrofurantoin, trimethoprim, or mecillinam. Non-S denotes resistant (R) or susceptible with increased exposure (I).*

***Supplementary Figure S2. Prediction performance for individual antibiotics using non-conformal prediction.*** *(A) Percentage of correct predictions for each antibiotic using AST results for four to eight antibiotics as input. (B) Distribution of prediction outcomes for each antibiotic. Green indicates correct predictions, yellow major errors (false-resistant predictions), and red very major errors (false-susceptible predictions). Percentages are calculated relative to all predictions. AMP, ampicillin; AMC, amoxicillin/clavulanic acid; PIP, piperacillin; TZP, piperacillin/tazobactam; CAZ, ceftazidime; CRO, ceftriaxone; CTX, cefotaxime; FEP, cefepime; CIP, ciprofloxacin; OFX, ofloxacin; LVX, levofloxacin; MFX, moxifloxacin; GEN, gentamicin; TOB, tobramycin; VME, very major errors; ME, major errors.*

***Supplementary Figure S3. Prediction performance for individual antibiotics with and without conformal prediction.*** *STD denotes standard (non-conformal) prediction. Conformal prediction was evaluated at confidence levels of 90%, 95%, and 97.5%, respectively. (A) Percentage of correct unambiguous predictions for each antibiotic. Only predictions assigned a single susceptibility category (S or R) were considered unambiguous. (B) Distribution of prediction outcomes for each antibiotic. Green indicates correct unambiguous predictions, yellow major errors (false-resistant predictions), and red very major errors (false-susceptible predictions). Percentages are calculated relative to all candidate predictions, including cases where the model abstained from prediction because the required confidence level was not achieved. AMP, ampicillin; AMC, amoxicillin/clavulanic acid; PIP, piperacillin; TZP, piperacillin/tazobactam; CAZ, ceftazidime; CRO, ceftriaxone; CTX, cefotaxime; FEP, cefepime; CIP, ciprofloxacin; OFX, ofloxacin; LVX, levofloxacin; MFX, moxifloxacin; GEN, gentamicin; TOB, tobramycin; VME, very major errors; ME, major errors.*

***Supplementary Figure S4. Performance metrics for individual antibiotics at different conformal prediction confidence levels using AST results for six antibiotics as input.*** *Performance was assessed using (A) Matthews correlation coefficient (MCC), (B) F1 score, (C) very major error (VME) rate, and (D) major error (ME) rate. Results are shown for standard (STD) prediction and conformal prediction at confidence levels of 90%, 95%, and 97.5%. AMP, ampicillin; AMC, amoxicillin/clavulanic acid; PIP, piperacillin; TZP, piperacillin/tazobactam; CAZ, ceftazidime; CRO, ceftriaxone; CTX, cefotaxime; FEP, cefepime; CIP, ciprofloxacin; OFX, ofloxacin; LVX, levofloxacin; MFX, moxifloxacin; GEN, gentamicin; TOB, tobramycin.*

***Supplementary Figure S5. Performance metrics by antibiotic class using non-conformal prediction and AST results for four to eight antibiotics as input.*** *Performance was assessed using (A) Matthews correlation coefficient (MCC), (B) F1 score, (C) very major error (VME) rate, and (D) major error (ME) rate. Antibiotics were grouped into penicillins, cephalosporins, fluoroquinolones, and aminoglycosides.*

***Supplementary Figure S6. Performance metrics by antibiotic class at different conformal prediction confidence levels using AST results for six antibiotics as input.*** *Performance was assessed using (A) Matthews correlation coefficient (MCC), (B) F1 score, (C) very major error (VME) rate, and (D) major error (ME) rate. Results are shown for standard (STD) prediction and conformal prediction at confidence levels of 90%, 95%, and 97.5%. Antibiotics were grouped into penicillins, cephalosporins, fluoroquinolones, and aminoglycosides.*

***Supplementary Figure S7. Performance metrics for input combinations constrained to include AST results for at least one antibiotic from each antibiotic class.*** *Performance was assessed using (A) Matthews correlation coefficient (MCC), (B) F1 score, (C) very major error (VME) rate, and (D) major error (ME) rate. Antibiotic classes were penicillins, cephalosporins, fluoroquinolones, and aminoglycosides. Results are shown for predictions using AST results for four to eight antibiotics as input.*

***Supplementary Figure S8. Performance metrics at different conformal prediction confidence levels using AST results for a fixed set of six antibiotics as input.*** *Performance was assessed using (A) Matthews correlation coefficient (MCC), (B) F1 score, (C) very major error (VME) rate, and (D) major error (ME) rate. Susceptibility to amoxicillin/clavulanic acid (AMC), piperacillin/tazobactam (TZP), cefotaxime (CTX), ciprofloxacin (CIP), ofloxacin (OFX), and tobramycin (TOB) was used to predict susceptibility to the remaining antibiotics. Results are shown for standard (STD) prediction and conformal prediction at confidence levels of 90%, 95%, and 97.5%.*

***Supplementary Figure S9. Performance metrics under alternative susceptibility category translation modes.*** *Performance was assessed using (A) Matthews correlation coefficient (MCC), (B) F1 score, (C) very major error (VME) rate, and (D) major error (ME) rate. Results are shown for predictions using AST results for six antibiotics as input. In Mode A, EUCAST categories S and I were treated as susceptible; in Mode B, category I was treated as resistant; and in Mode C, both category I and measurements within the area of technical uncertainty (ATU) were treated as resistant. AMP, ampicillin; AMC, amoxicillin/clavulanic acid; PIP, piperacillin; TZP, piperacillin/tazobactam; CAZ, ceftazidime; CRO, ceftriaxone; CTX,* *cefotaxime; FEP, cefepime; CIP, ciprofloxacin; OFX, ofloxacin; LVX, levofloxacin; MFX, moxifloxacin; GEN, gentamicin; TOB, tobramycin.*

***Supplementary Figure S10. Resistance gene groups in relation to inhibition zone diameters.*** *(A) Quinolone resistance determinants. (B) Aminoglycoside resistance determinants. (C) Beta-lactamase groups. Rows represent individual isolates ordered according to hierarchical clustering of inhibition zone diameters. Heatmap intensity reflects inhibition zone diameter, ranging from maximum inhibition (white) to no visible inhibition zone (dark red). Side annotations indicate phenotype cluster assignment and resistance gene group presence. AMP, ampicillin; AMC, amoxicillin/clavulanic acid; PIP, piperacillin; TZP, piperacillin/tazobactam; CAZ, ceftazidime; CRO, ceftriaxone; CTX, cefotaxime; FEP, cefepime; CIP, ciprofloxacin; OFX, ofloxacin; LVX, levofloxacin; MFX, moxifloxacin; GEN, gentamicin; TOB, tobramycin. QRDR, quinolone resistance-determining region mutations; PMQR, plasmid-mediated quinolone resistance genes; AAC, aminoglycoside N-acetyltransferases; APH, aminoglycoside O-phosphotransferases; AAD, aminoglycoside adenyltransferases; ANT, aminoglycoside nucleotidyltransferases; ESBL carba, carbapenemases; AmpC, AmpC beta-lactamases; ESBL classic, extended-spectrum beta-lactamases excluding AmpC beta-lactamases and carbapenemases; Non-ESBL, beta-lactamases without extended-spectrum activity.*

***Supplementary Figure S11. Resistance gene symbols in relation to inhibition zone diameters and susceptibility phenotypes.*** *(A) Quinolone resistance gene symbols and mutations. (B) Aminoglycoside resistance gene symbols. (C) Beta-lactamase gene symbols. (D) EUCAST susceptibility categories. Rows represent individual isolates ordered according to hierarchical clustering of inhibition zone diameters. Heatmap intensity reflects inhibition zone diameter, ranging from maximum inhibition (white) to no visible inhibition zone (dark red). The EUCAST susceptibility panel is annotated with phenotype cluster assignment; red indicates resistant (R), yellow susceptible with increased exposure (I), and green susceptible (S). Asterisks indicate measurements within the area of technical uncertainty (ATU). AMP, ampicillin; AMC, amoxicillin/clavulanic acid; PIP, piperacillin; TZP, piperacillin/tazobactam; CAZ, ceftazidime; CRO, ceftriaxone; CTX, cefotaxime; FEP, cefepime; CIP, ciprofloxacin; OFX, ofloxacin; LVX, levofloxacin; MFX, moxifloxacin; GEN, gentamicin; TOB, tobramycin. Gene symbols are defined according to the NCBI Reference Gene Catalog.*

***Supplementary Figure S12. Isolate-level mean absolute prediction error and resistance determinant groups.*** *(A) Heatmap of EUCAST susceptibility categories for isolates ordered by isolate-level mean absolute error. The EUCAST susceptibility panel is annotated with phenotype cluster assignment; red indicates resistant (R), yellow susceptible with increased exposure (I), and green susceptible (S). Asterisks indicate measurements within the area of technical uncertainty (ATU). (B) Distribution of isolate-level mean absolute error according to resistance determinant group. Each point represents one isolate. Boxplots show distributions of isolate-level mean absolute error stratified by the presence or absence of resistance determinants belonging to each resistance gene group. The center line indicates the median, boxes represent the interquartile range (IQR), whiskers extend to 1.5 × IQR, and points represent individual isolates outside this range. Mean absolute error was calculated across all antibiotics and evaluated combinations of AST results for six antibiotics as input. AMP, ampicillin; AMC, amoxicillin/clavulanic acid; PIP, piperacillin; TZP, piperacillin/tazobactam; CAZ, ceftazidime; CRO, ceftriaxone; CTX, cefotaxime; FEP, cefepime; CIP, ciprofloxacin; OFX, ofloxacin; LVX, levofloxacin; MFX, moxifloxacin; GEN, gentamicin; TOB, tobramycin; VME, very major errors; ME, major errors. QRDR, quinolone resistance-determining region mutations; PMQR, plasmid-mediated quinolone resistance genes; AAC, aminoglycoside N-acetyltransferases; APH, aminoglycoside O-phosphotransferases; AAD, aminoglycoside adenyltransferases; ANT, aminoglycoside nucleotidyltransferases; ESBL carba, carbapenemases; AmpC, AmpC beta-lactamases; ESBL classic, extended-spectrum beta-lactamases excluding AmpC beta-lactamases and carbapenemases; Non-ESBL, beta-lactamases without extended-spectrum activity.*


1. Ward Jr JH. 1963. Hierarchical grouping to optimize an objective function. Journal of the American statistical association 58:236–244.
2. Rousseeuw PJ. 1987. Silhouettes: a graphical aid to the interpretation and validation of cluster analysis. Journal of computational and applied mathematics 20:53–65.
3. Marmur J. 1961. A procedure for the isolation of deoxyribonucleic acid from micro-organisms. Journal of Molecular Biology 3:208–IN1.
4. Salvà-Serra F, Gomila M, Svensson-Stadler L, Busquets A, Jaén-Luchoro D, Karlsson R, Moore ER. 2018. A protocol for extraction and purification of high-quality and quantity bacterial DNA applicable for genome sequencing: a modified version of the Marmur procedure. doi:10.1038/protex.2018.084.

5. Sanger F, Nicklen S, Coulson AR. 1977. DNA sequencing with chain-terminating inhibitors. Proceedings of the national academy of sciences 74:5463–5467.
6. Jaén-Luchoro D, Al-Shaer S, Piñeiro-Iglesias B, Gonzales-Siles L, Cardew S, Jensie-Markopolous S, Ohlén M, Inganäs E, Neumann-Schaal M, Wolf J. 2023. Corynebacterium genitalium sp. nov., nom. rev. and Corynebacterium pseudogenitalium sp. nov., nom. rev., two old species of the genus Corynebacterium described from clinical and environmental samples. Research in Microbiology 174:103987.
7. Lane D. 1991. 16S/23S rRNA sequencing. Nucleic acid techniques in bacterial systematics.
8. Hauben L, Vauterin L, Swings J, Moore E. 1997. Comparison of 16S ribosomal DNA sequences of all Xanthomonas species. International Journal of Systematic and Evolutionary Microbiology 47:328–335.
9. Fredricks DN, Relman DA. 1998. Improved amplification of microbial DNA from blood cultures by removal of the PCR inhibitor sodium polyanetholesulfonate. Journal of clinical microbiology 36:2810–2816.
10. Low AJ, Koziol AG, Manninger PA, Blais B, Carrillo CD. 2019. ConFindr: rapid detection of intraspecies and cross-species contamination in bacterial whole-genome sequence data. PeerJ 7:e6995.
11. Krueger F. 2015. Trim Galore!: A wrapper around Cutadapt and FastQC to consistently apply adapter and quality trimming to FastQ files, with extra functionality for RRBS data. Babraham Institute.
12. Martin M. 2011. Cutadapt removes adapter sequences from high-throughput sequencing reads. EMBnet journal 17:10–12.
13. Andrews S. 2010. FastQC: a quality control tool for high throughput sequence data. Babraham Bioinformatics, Babraham Institute, Cambridge, United Kingdom.
14. Prjibelski A, Antipov D, Meleshko D, Lapidus A, Korobeynikov A. 2020. Using SPAdes de novo assembler. Current protocols in bioinformatics 70:e102.
15. Mikheenko A, Prjibelski A, Saveliev V, Antipov D, Gurevich A. 2018. Versatile genome assembly evaluation with QUAST-LG. Bioinformatics 34:i142–i150.
16. Parks DH, Imelfort M, Skennerton CT, Hugenholtz P, Tyson GW. 2015. CheckM: assessing the quality of microbial genomes recovered from isolates, single cells, and metagenomes. Genome research 25:1043–1055.
17. Richter M, Rosselló-Móra R, Oliver Glöckner F, Peplies J. 2016. JSpeciesWS: a web server for prokaryotic species circumscription based on pairwise genome comparison. Bioinformatics 32:929–931.
18. Meier-Kolthoff JP, Göker M. 2019. TYGS is an automated high-throughput platform for state-of-the-art genome-based taxonomy. Nature communications 10:2182.
19. Ewels P, Magnusson M, Lundin S, Käller M. 2016. MultiQC: summarize analysis results for multiple tools and samples in a single report. Bioinformatics 32:3047–3048.
20. Wirth T, Falush D, Lan R, Colles F, Mensa P, Wieler LH, Karch H, Reeves PR, Maiden MC, Ochman H, Achtman M. 2006. Sex and virulence in *Escherichia coli*: an evolutionary perspective. Mol Microbiol 60:1136–51.
21. Jolley KA, Bray JE, Maiden MC. 2018. Open-access bacterial population genomics: BIGSdb software, the PubMLST. org website and their applications. Wellcome open research 3:124.
22. Dyer NP, Päuker B, Baxter L, Gupta A, Bunk B, Overmann J, Diricks M, Dreyer V, Niemann S, Holt KE. 2025. EnteroBase in 2025: exploring the genomic epidemiology of bacterial pathogens. Nucleic acids research 53:D757–D762.
23. Feldgarden M, Brover V, Gonzalez-Escalona N, Frye JG, Haendiges J, Haft DH, Hoffmann M, Pettengill JB, Prasad AB, Tillman GE. 2021. AMRFinderPlus and the Reference Gene Catalog facilitate examination of the genomic links among antimicrobial resistance, stress response, and virulence. Scientific reports 11:12728.

24. Hunt M, Mather AE, Sánchez-Busó L, Page AJ, Parkhill J, Keane JA, Harris SR. 2017. ARIBA: rapid antimicrobial resistance genotyping directly from sequencing reads. Microbial genomics 3:e000131.
25. Zankari E, Hasman H, Cosentino S, Vestergaard M, Rasmussen S, Lund O, Aarestrup FM, Larsen MV. 2012. Identification of acquired antimicrobial resistance genes. Journal of antimicrobial chemotherapy 67:2640–2644.
26. Bush K, Jacoby GA. 2010. Updated functional classification of β-lactamases. Antimicrobial agents and chemotherapy 54:969–976.
27. Naas T, Oueslati S, Bonnin RA, Dabos ML, Zavala A, Dortet L, Retailleau P, Iorga BI. 2017. Beta-lactamase database (BLDB)–structure and function. Journal of enzyme inhibition and medicinal chemistry 32:917–919.
