## Supplementary figures S1-S6 for "Clinical evaluation of artificial intelligence for diagnostics of antibiotic-resistant bacteria"

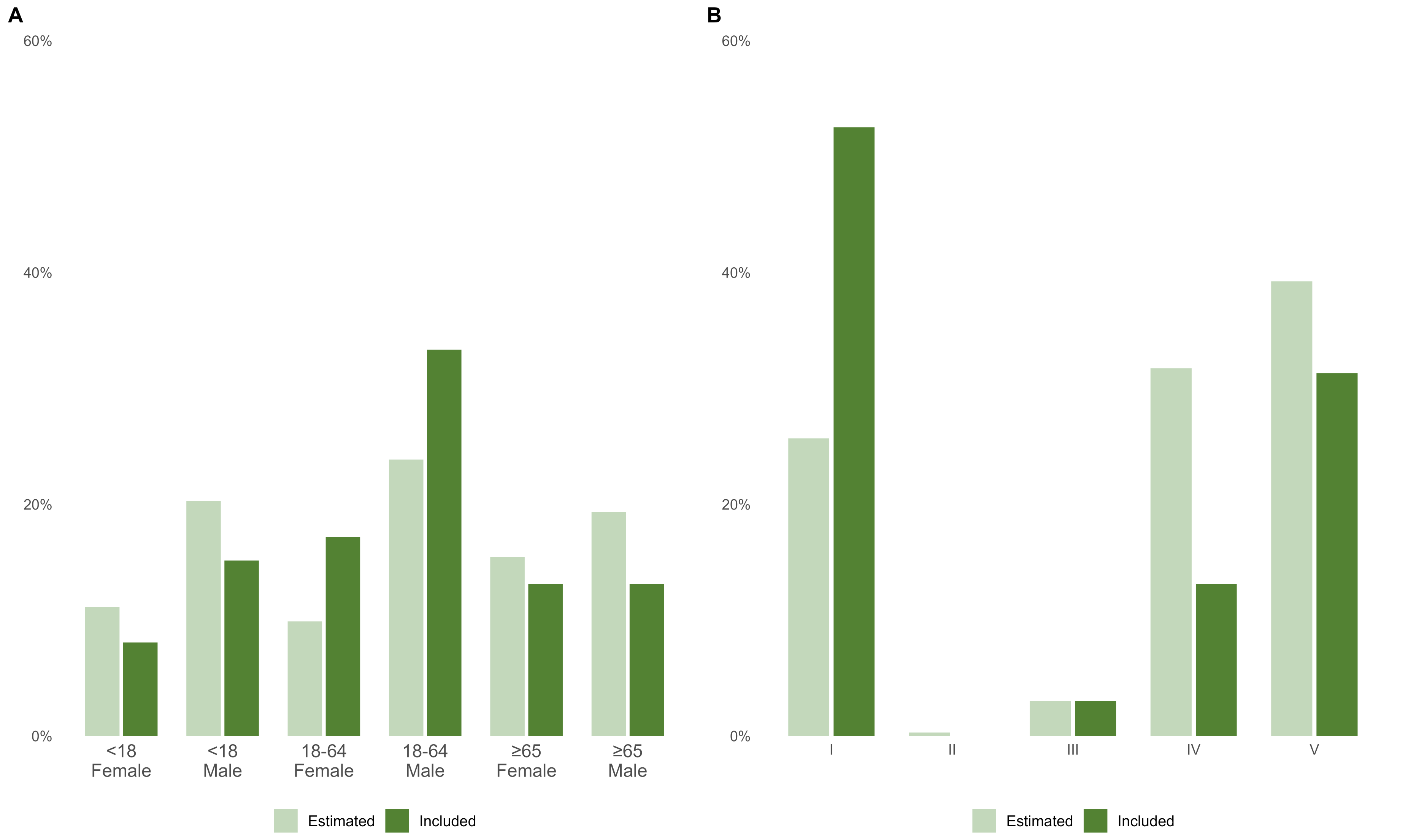


**Supplementary Figure S1. Expected and observed distributions of included isolates.**

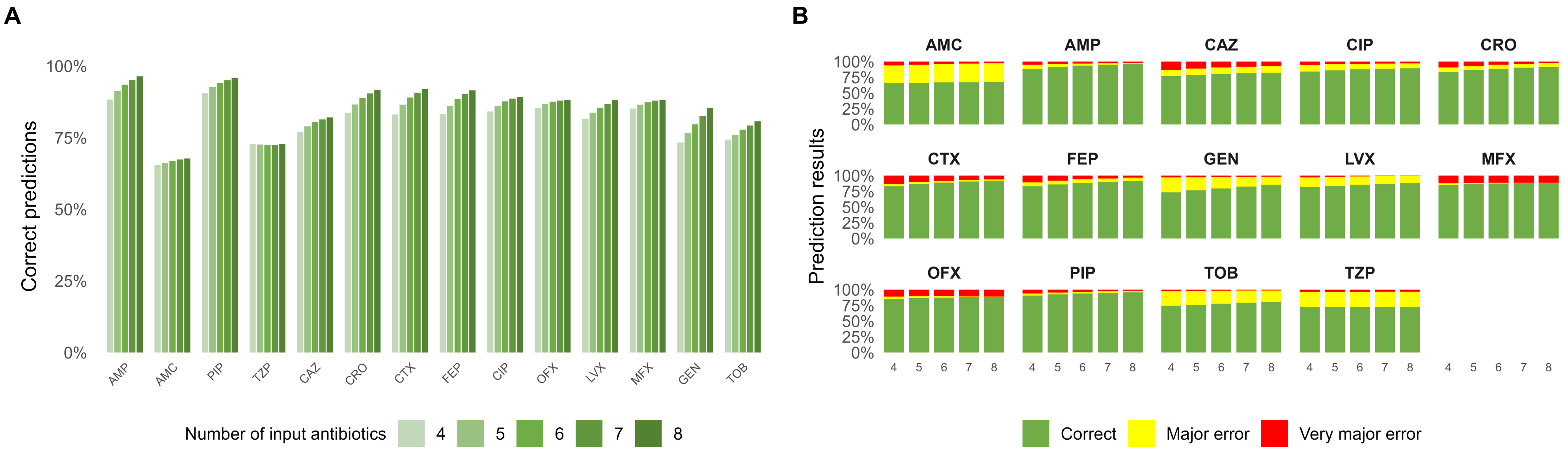


**Supplementary Figure S2. Prediction performance for individual antibiotics using non−conformal prediction.**

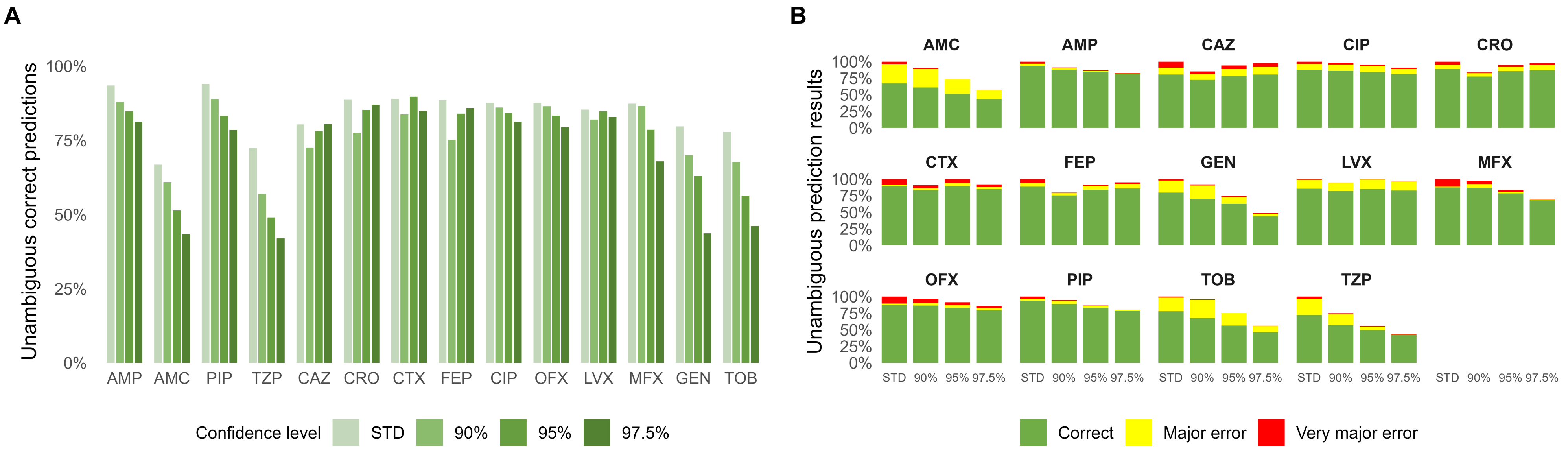


**Supplementary Figure S3. Prediction performance for individual antibiotics with and without conformal prediction.**

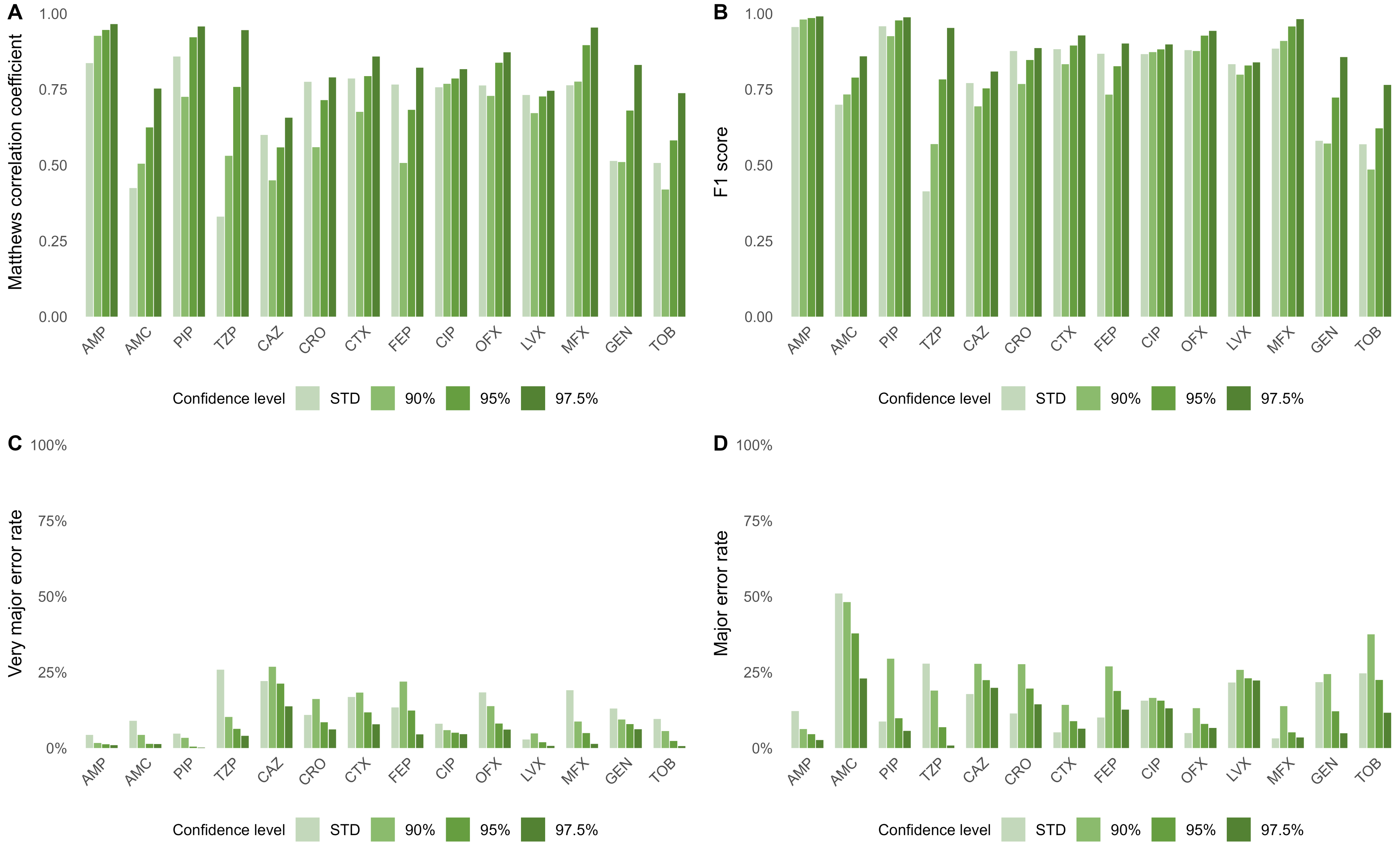


**Supplementary Figure S4. Performance metrics for individual antibiotics at different conformal prediction confidence levels using AST results for six antibiotics as input.**

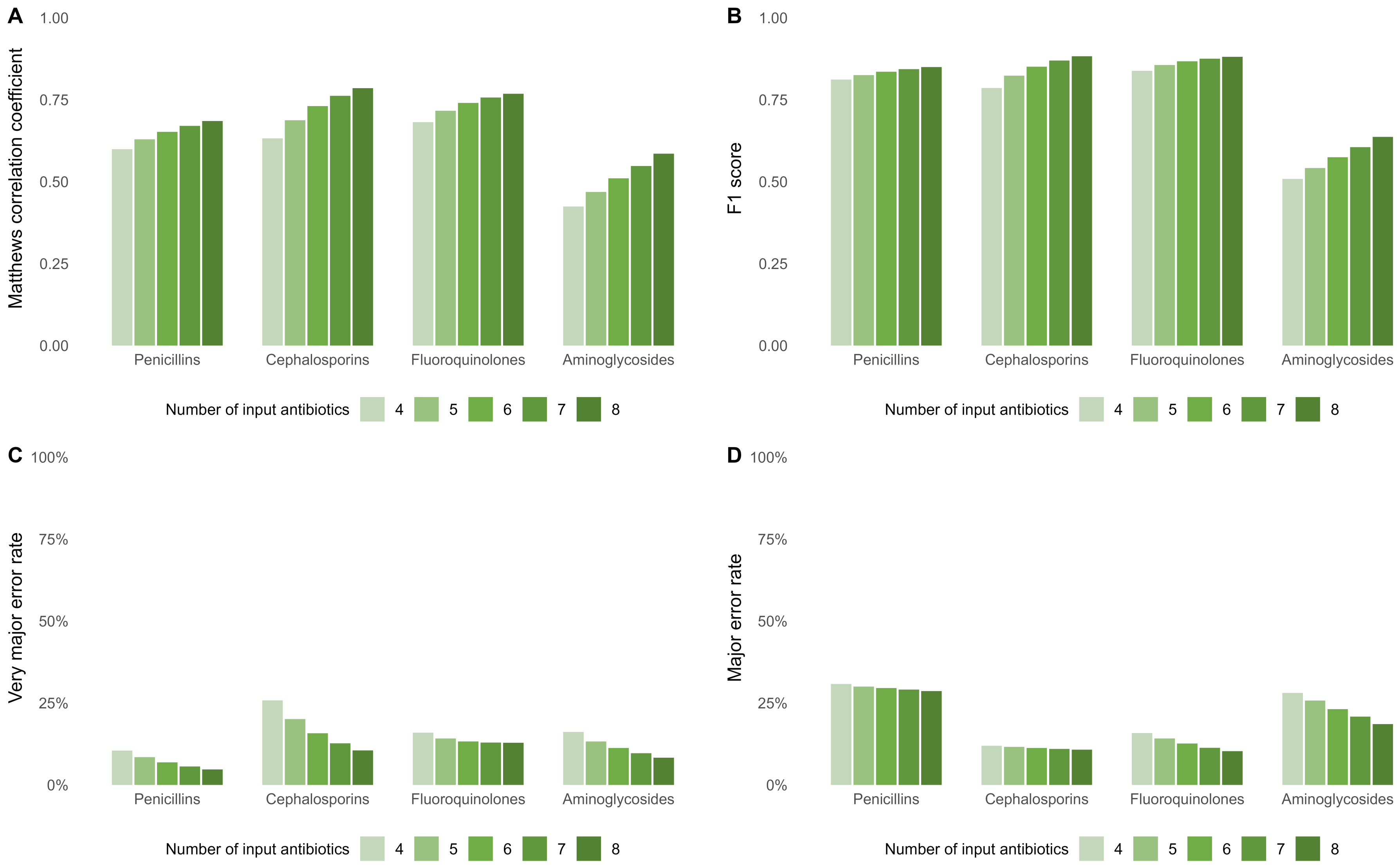


**Supplementary Figure S5. Performance metrics by antibiotic class using non−conformal prediction and AST results for four to eight antibiotics as input.**

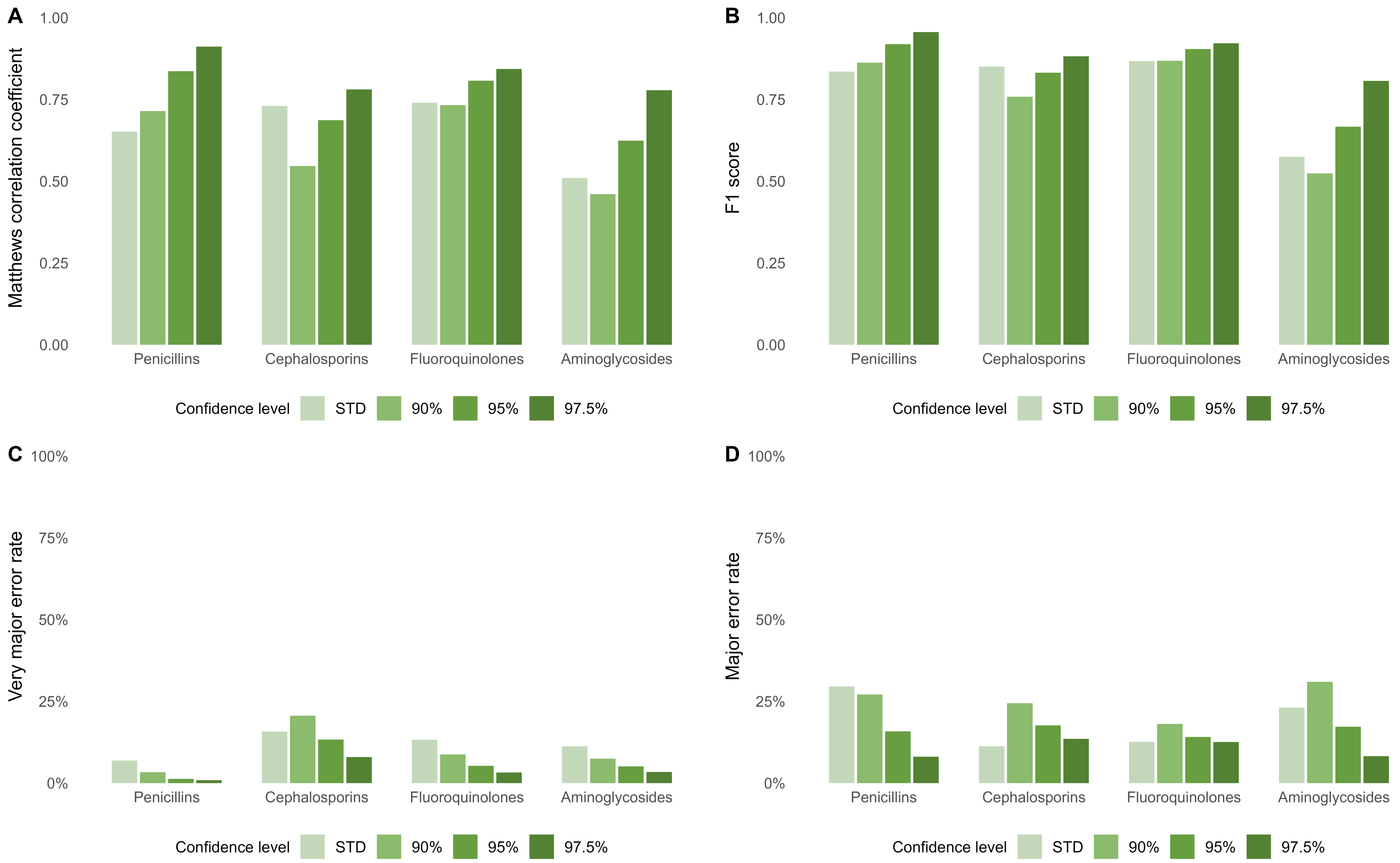


**Supplementary Figure S6. Performance metrics by antibiotic class at different conformal prediction confidence levels using AST results for six antibiotics as input.**
