## Supplementary figures S7-S12 for "Clinical evaluation of artificial intelligence for diagnostics of antibiotic-resistant bacteria"

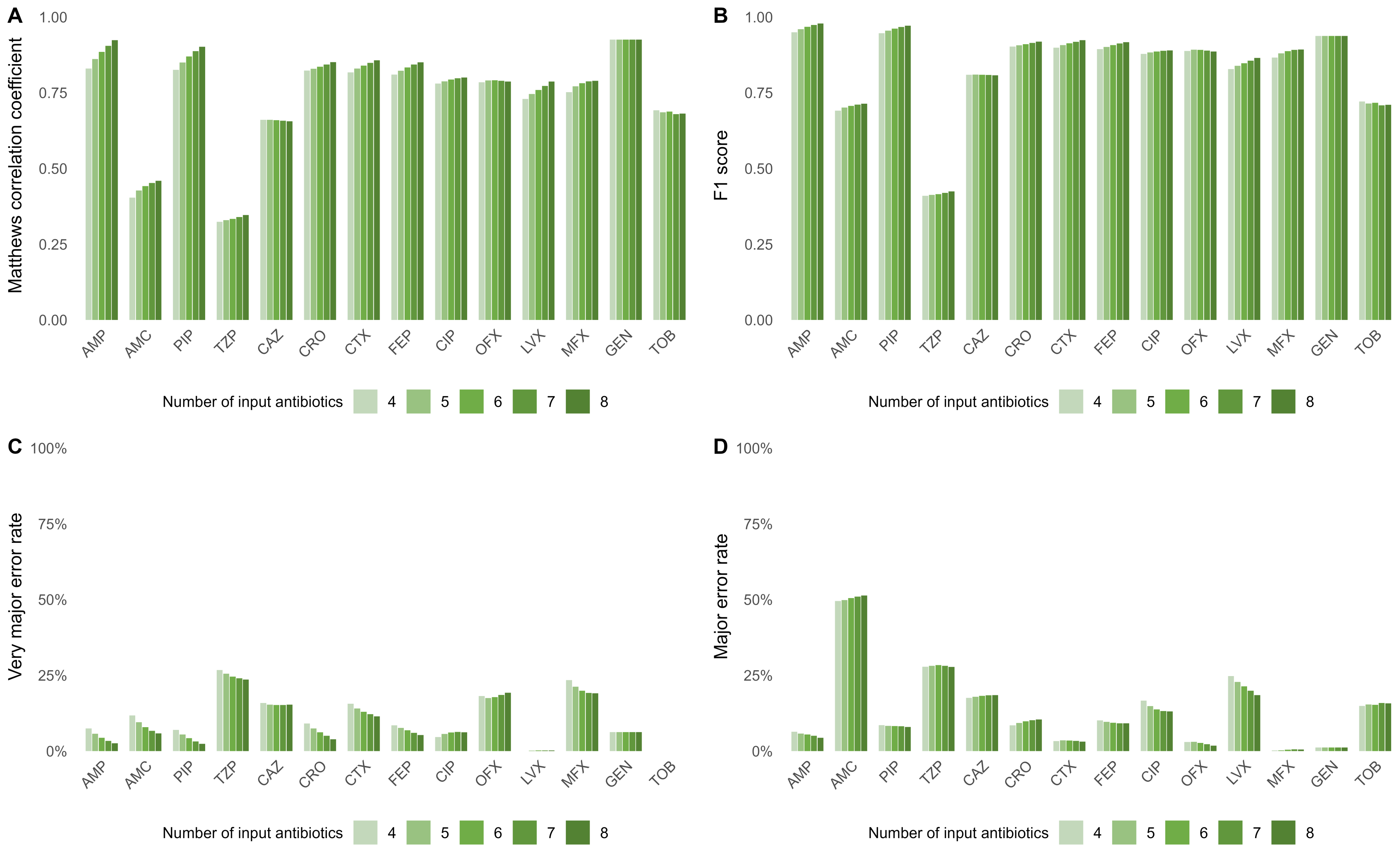


**Supplementary Figure S7. Performance metrics for input combinations constrained to include AST results for at least one antibiotic from each antibiotic class.**

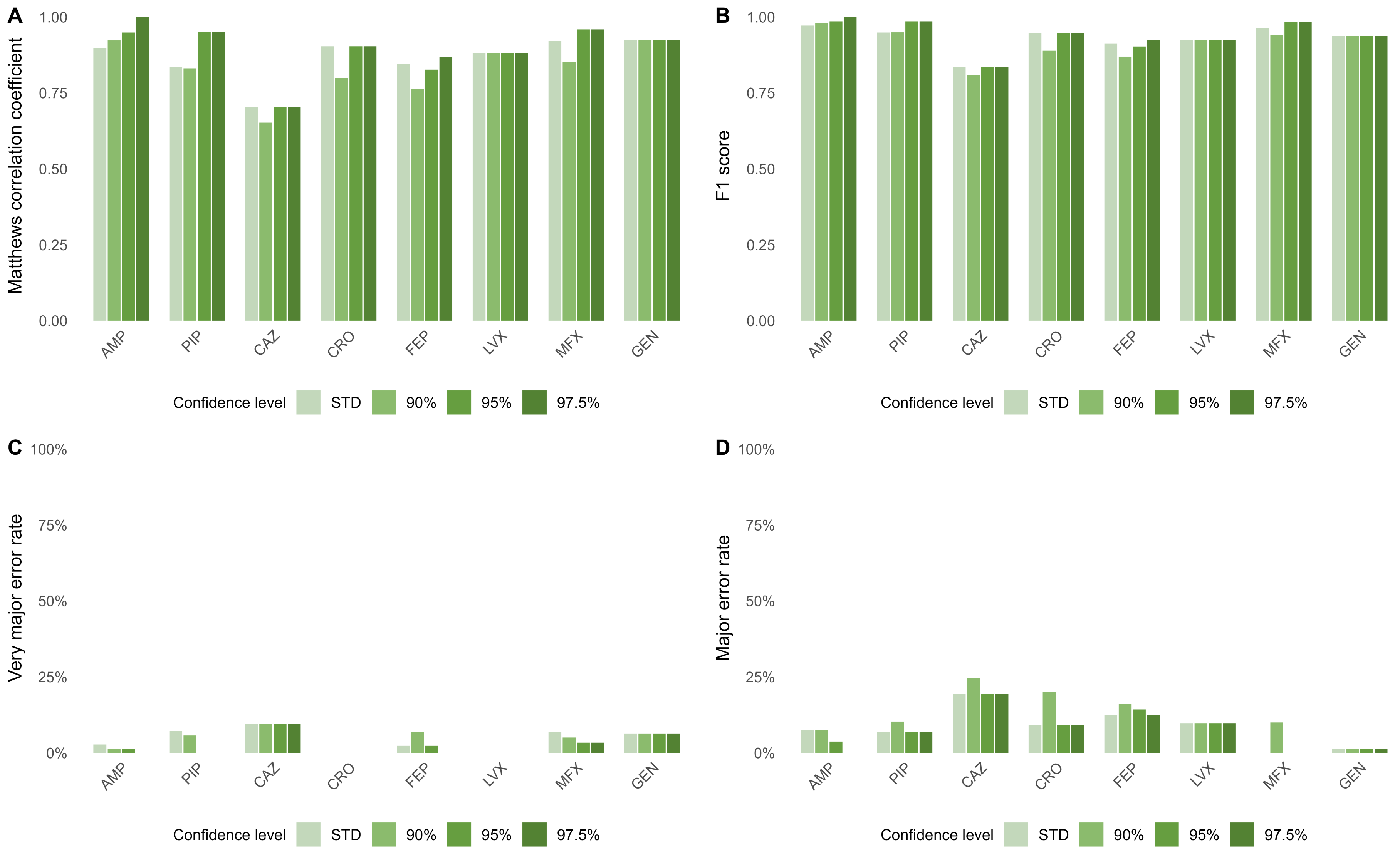


**Supplementary Figure S8. Performance metrics at different conformal prediction confidence levels using AST results for a fixed set of six antibiotics as input.**

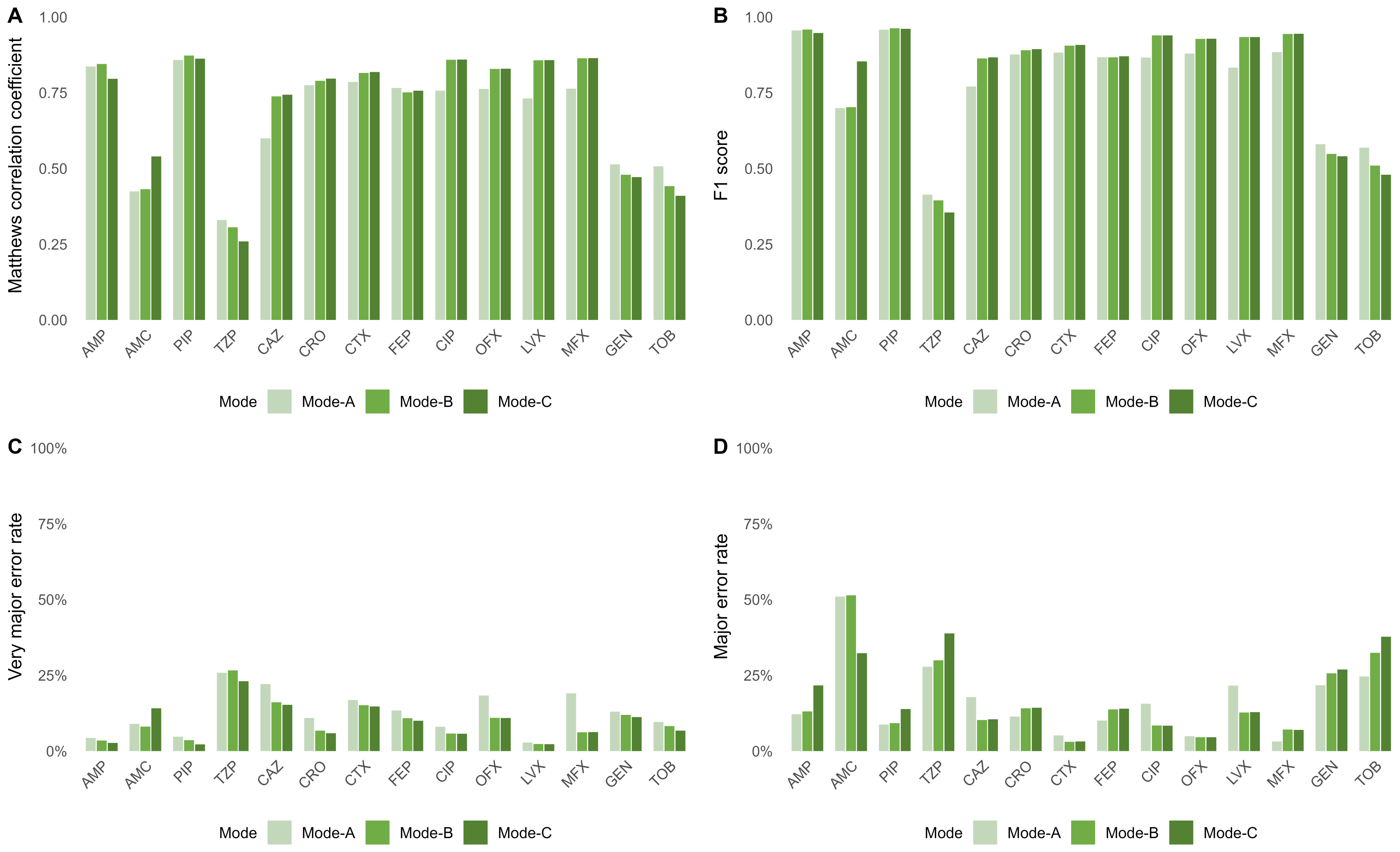


**Supplementary Figure S9. Performance metrics under alternative susceptibility category translation modes.**

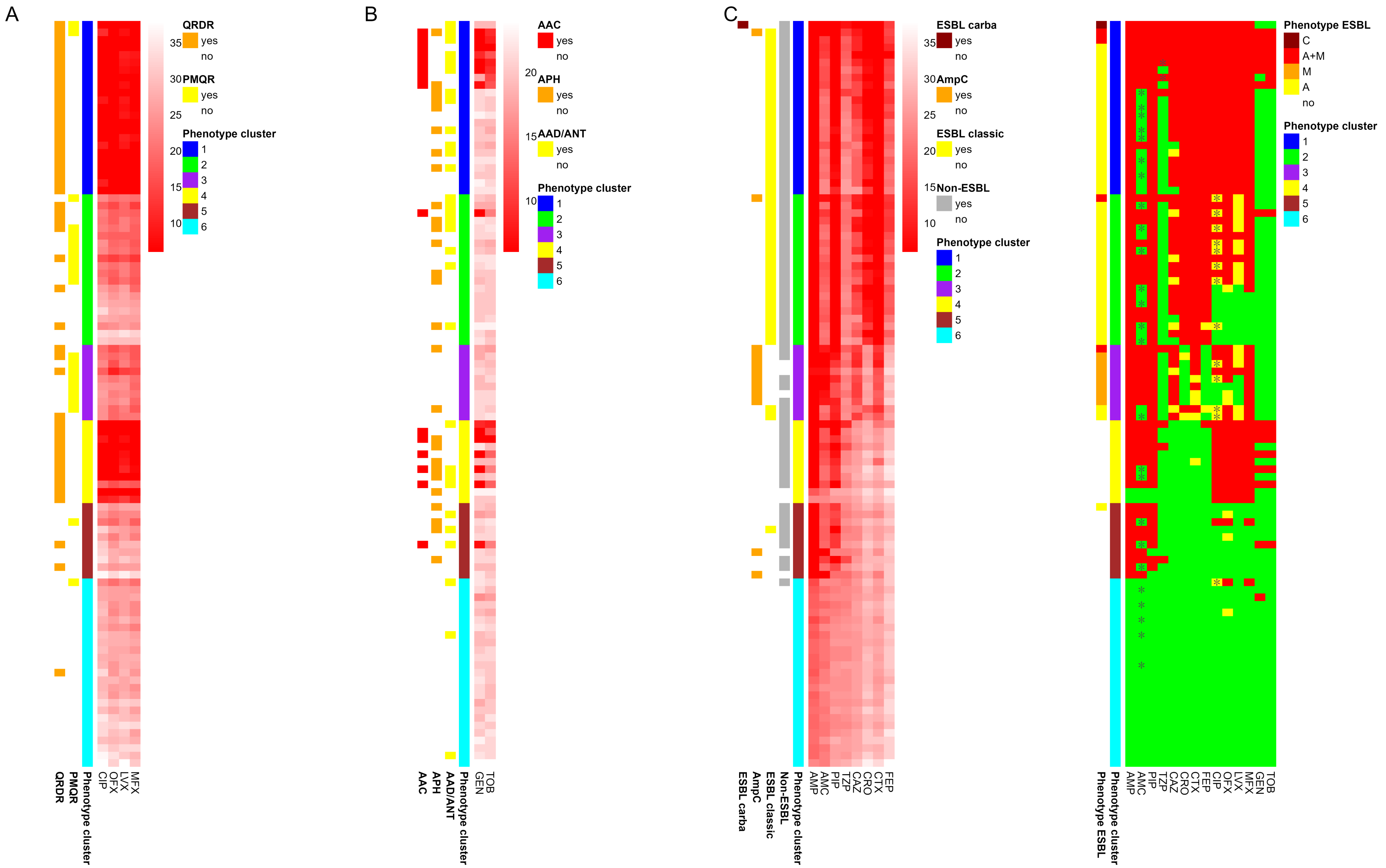


**Supplementary Figure S10. Resistance gene groups in relation to inhibition zone diameters.**

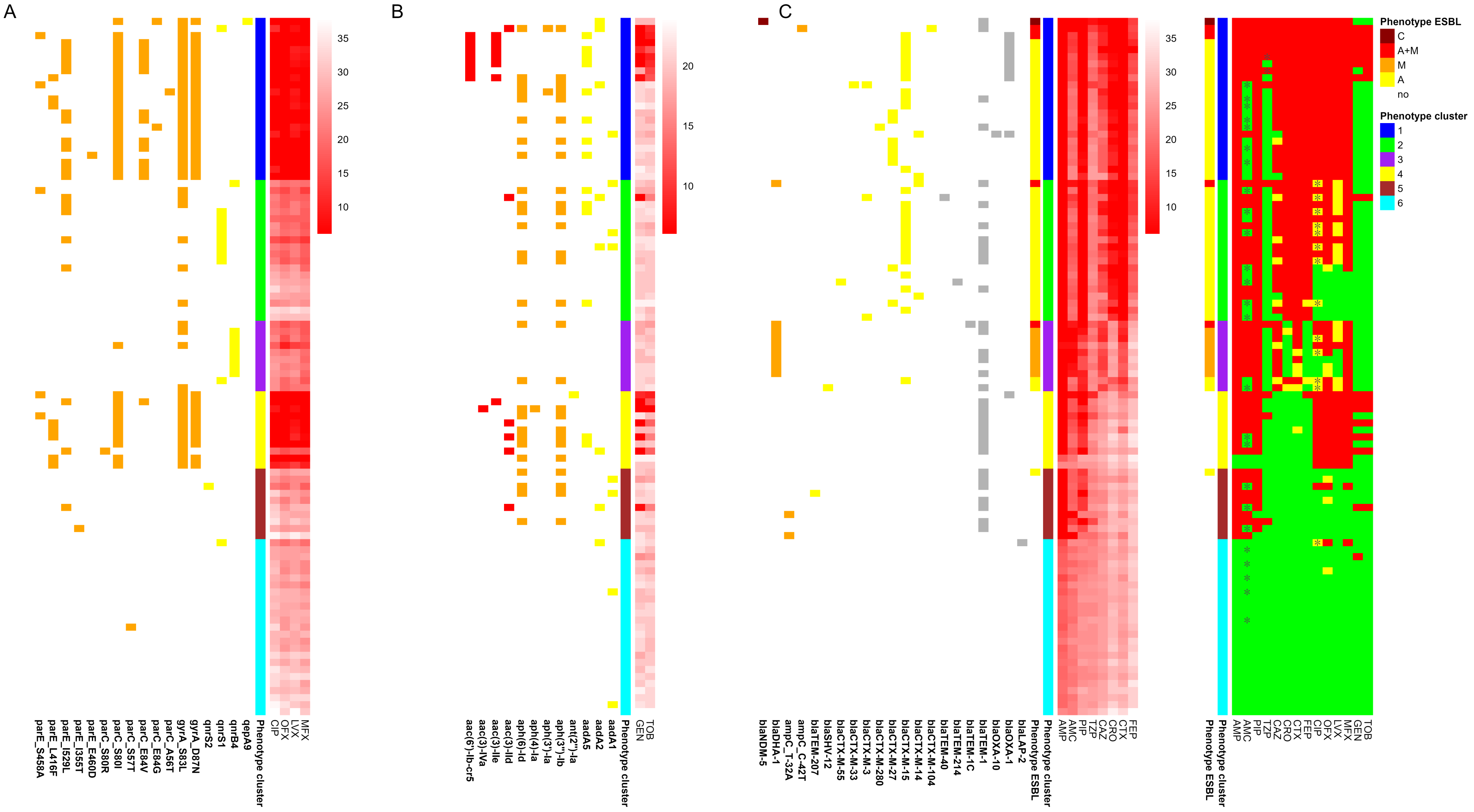


**Supplementary Figure S11. Resistance gene symbols in relation to inhibition zone diameters and susceptibility phenotypes.**

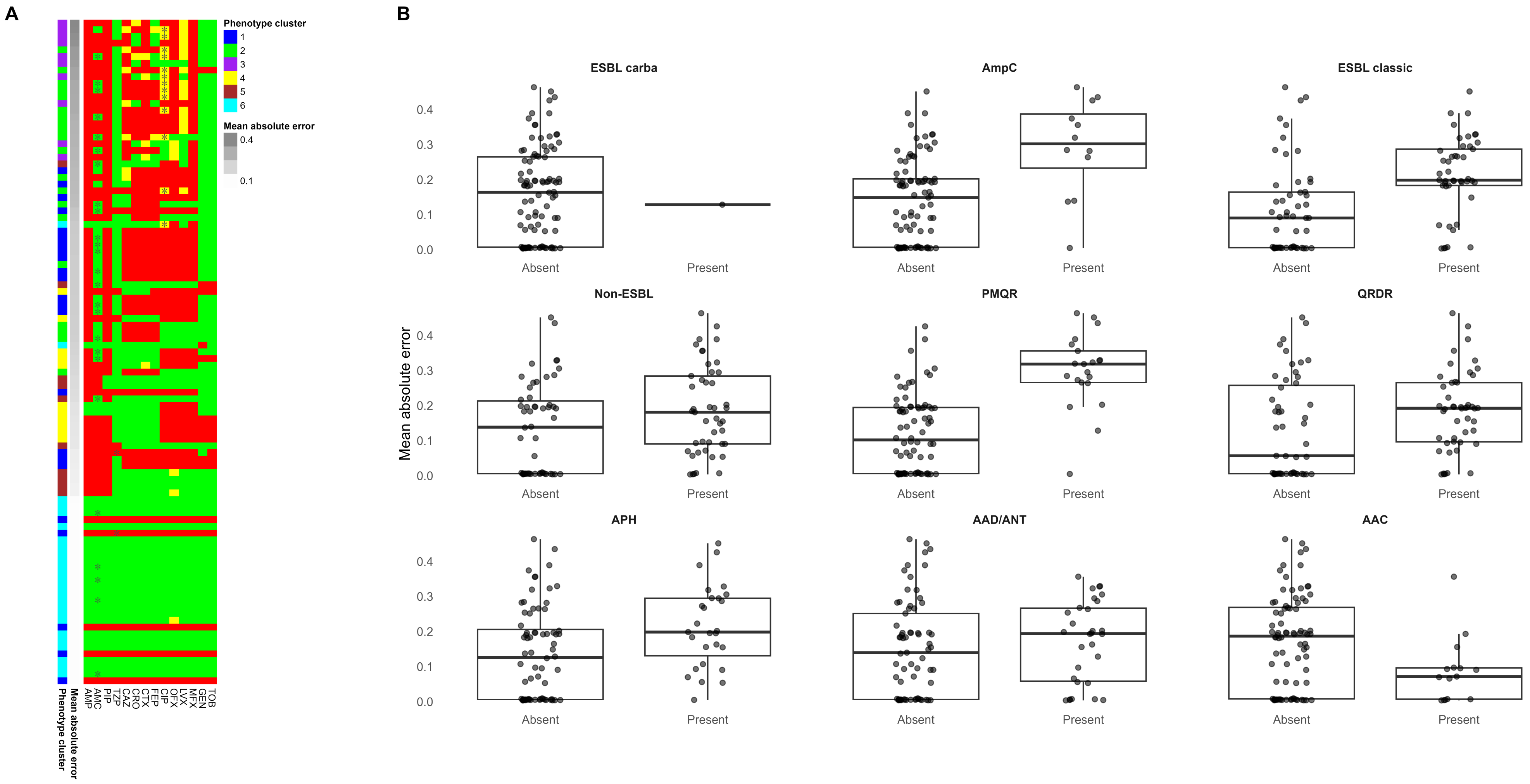


**Supplementary Figure S12. Isolate−level mean absolute prediction error and resistance determinant groups.**
